# Phylogeography and transmission of *M. tuberculosis* in Moldova

**DOI:** 10.1101/2021.06.30.21259748

**Authors:** Chongguang Yang, Benjamin Sobkowiak, Vijay Naidu, Alexandru Codreanu, Nelly Ciobanu, Kenneth S. Gunasekera, Melanie H. Chitwood, Sofia Alexandru, Stela Bivol, Marcus Russi, Joshua Havumaki, Patrick Cudahy, Heather Fosburgh, Christopher J. Allender, Heather Centner, David M. Engelthaler, Nicolas A. Menzies, Joshua L. Warren, Valeriu Crudu, Caroline Colijn, Ted Cohen

## Abstract

**Background:** The incidence of multidrug-resistant tuberculosis (MDR-TB) remains critically high in countries of the former Soviet Union, where >20% of new cases and >50% of previously-treated cases have resistance to rifampin and isoniazid. Transmission of resistant strains, as opposed to resistance selected through inadequate treatment of drug-susceptible TB, is the main driver of incident MDR-TB in these countries.

**Methods:** We conducted a prospective, genomic analysis of all culture-positive TB cases diagnosed in 2018 and 2019 in the Republic of Moldova. We used phylogenetic methods to identify putative transmission clusters; spatial and demographic data were analyzed to further describe local transmission of *M. tuberculosis*.

**Results:** Of 2236 participants, 779 (36%) had MDR-TB, of whom 386 (50%) had never been treated previously for TB. 92% of MDR *M. tuberculosis* strains belonged to putative transmission clusters. Phylogenetic reconstruction identified three large clades that were comprised nearly uniformly of MDR-TB; two of these clades were of Beijing lineage and one of Ural lineage, and each had additional distinct clade-specific second-line drug resistance mutations and geographic distributions. Spatial and temporal proximity between pairs of cases within a cluster was associated with greater genomic similarity.

**Conclusions:** The MDR-TB epidemic in Moldova is the result of local transmission of multiple *M. tuberculosis* strains, including distinct clades of highly drug-resistant *M. tuberculosis* with varying geographic distributions and drug resistance profiles. This study demonstrates the role of comprehensive genomic surveillance for understanding the transmission of *M. tuberculosis* and highlights the urgency of interventions to interrupt transmission of highly drug-resistant *M. tuberculosis*.

## Introduction

Multidrug-resistant tuberculosis (MDR-TB) (i.e., resistance to at least rifampin and isoniazid) poses serious threats to effective TB control in many countries. Globally, approximately 4-5% of incident TB cases are MDR, but this is substantially higher in countries of the former Soviet Union where MDR-TB represents >20% of new TB cases and >50% of previously-treated TB.^1^ MDR-TB in this region has been attributed to breakdowns in public health infrastructure, transmission of TB in hospitals and prisons, and a deterioration of living conditions coinciding with the dissolution of the Soviet Union in the early 1990s.^2^ While the contributions of these factors remain uncertain, there is consensus that the transmission of MDR-TB, as opposed to resistance acquired through inadequate treatment of drug susceptible TB, is now the predominant cause of incident MDR-TB.^3^ This consensus is supported by routine surveillance data that document that the majority of incident MDR-TB episodes are diagnosed among individuals with no prior anti-TB treatment.^1^ However, these data alone do not address critical questions about where and between whom MDR-TB is transmitted or reveal the extent to which specific *M. tuberculosis* variants are responsible for MDR-TB transmission.

The increasing availability of next generation sequencing (NGS), coupled with the development of analytic approaches for integrating high-resolution genomic, spatial, and epidemiological data, has transformed our ability to describe transmission of pathogens in populations.^4-6^ Previous genomic analyses of TB from the former Soviet Union have described the emergence and evolution of specific *M. tuberculosis* lineages responsible for an outsized proportion of MDR-TB in the region. In general, these studies have been conducted on isolates enriched for drug resistance phenotypes or on samples from larger cohorts ^7,8,9^, and this can challenge transmission inference.

We systematically collected and sequenced initial diagnostic isolates from all culture-positive TB cases occurring over two years in the Republic of Moldova, a former Soviet country experiencing a severe MDR-TB epidemic. In addition to capturing *M. tuberculosis* isolates from all culture-positive cases, we also collected data on home location and other demographic and epidemiological data, allowing us to study the distribution and dynamics of TB with high resolution across the entire country.

## Methods

### Study setting

Moldova is a small country (∼ four million population), which gained independence when the Soviet Union dissolved in 1991. In 2019, the World Health Organization estimated an incidence rate of 80 TB cases (68-92) per 100,000 persons. 33% (30-35%) of new TB cases and 60% (56-64%) of previously treated TB cases were estimated to have MDR-TB.^1^

### Participant enrolment

TB diagnosis occurs at 46 diagnostic centers located throughout the country. Between Jan 1, 2018 and Dec 31, 2019, all non-incarcerated individuals evaluated for pulmonary TB were invited to participate in this study. Consenting participants allowed us to access clinical data and to perform sequencing on their mycobacterial isolates should they have culture-positive tuberculosis. This study was approved by the Ethics Committee of Research of the Phthisiopneumology Institute in Moldova and the Yale University Human Investigation Committee (No. 2000023071).

### Data and specimen collection and processing

Demographic data (sex, age, employment, history of incarceration, education level), residential status (rural or urban residence, home village/locality), and epidemiological data (household contacts, date of diagnosis) were collected from each participant.

Sputum specimens were tested at diagnostic centers by microscopy and Xpert and then sent to four in-country laboratories for solid and liquid culture. Positive cultures were sent to the National TB Reference Laboratory in Chisinau for mycobacterial DNA extraction by the cetyl trimethyl ammonium bromide (CTAB) method^10^.

### Whole-genome sequencing

Genomic DNA was prepared for NGS using the Illumina DNA Prep library preparation kit (supplementary appendix). Raw sequencing files were checked with FastQC^11^ and mapped to the H37Rv reference strain (NC_000962.3) using BWA ‘mem ‘^12^ and sorted with SAMtools v.1.10.^13^ Variant calling was conducted with GATK^14^ to identify single nucleotide polymorphisms (SNPs), with low-quality SNPs (Phred score Q < 20 and read depth < 5) and sites with missing calls in >10% of isolates removed.

Samples with possible polyclonal infections were identified through a previously detailed approach^15^ and removed from further analysis (Supplementary Appendix). Heterogenous sites were called as the consensus allele if present in ≥80% of mapped reads, otherwise they were labeled as ambiguous. SNPs in repetitive regions, PE/PPE genes and in known resistance-conferring genes were excluded from phylogenetic tree reconstruction. *In silico* drug resistance prediction was carried out using TB-Profiler v2.8.14.^16^

### Phylogenetic analysis and transmission cluster identification

A multiple sequence alignment of concatenated SNPs was used to construct a maximum-likelihood (M-L) phylogenetic tree with RAxML,^17^ using the ‘GTRGAMMA ‘ nucleotide substitution model and a Lewis ascertainment bias correction from 500 bootstrap samples. Putative transmission clusters were identified in the resulting M-L tree using TreeCluster^18^, testing two distance thresholds of 0·001 and 0·0005 substitutions/site, corresponding to approximate SNP thresholds of 40 and 20, respectively. Timed phylogenetic trees for each large cluster (≥10 cases) identified using the distance threshold of 0·001 substitutions/site were built with BEAST2 v2.6.3.^19^ (supplementary appendix). Briefly, phylogenetic trees were built using a strict molecular clock with a fixed rate of 1.0×10^−7^ per site per year and constant population model with a log normal [0,200] prior distribution.^20^ Markov chain Monte Carlo chains were run for 250 million iterations, with 10% burn-in to produce maximum clade credibility trees. Finally, past population events in three large clades identified in the study population were inferred using the Bayesian Skyline model in BEAST2.

### Spatial/genetic distance analysis

For each large transmission cluster (≥ten cases), we used multiple linear regression to quantify the association between the genetic and spatial distances for unique pairs of cases, adjusting for other pair- and individual-level features. We then used a Bayesian meta-analysis framework to better understand shared trends and variability in the estimated associations across genetic clusters. In our main analysis we present results using log-scaled patristic distances; as sensitivity analysis, we use SNP distances and a modified Poisson regression framework (details in Supplementary Appendix).

## Results

### Study population

We invited all culture-positive TB patients (N=2770) over the study period to participate; 2405 consented and among them, 2236 (93%) had available isolates for NGS analysis. These patients lived in 709 named localities within 50 regions (**Fig. 1**). Among enrolled participants with treatment history information (N=2182, **Table 1**), 31% had been previously treated for TB, 22% were female, and the median age was 43 years (interquartile range (IQR) 23-71). 60% lived in rural regions and 10% were previously imprisoned.

**Table 1.** Demographic and clinical characteristics of study participants.

| Characteristics | All participants<br>(N=2182*) | New cases<br>(N=1514) | Previously treated<br>cases (N=668) |
| --- | --- | --- | --- |
| **Demographic** |  |  |  |
| Female, no. (%) | 482 (22·1) | 350 (23·1) | 132 (19·8) |
| Age, Median (yr, IQR) | 43 (23-71) | 42 (27-65) | 43 (22-68) |
| Homeless | 238 (10·9) | 144 (9·5) | 94 (14·1) |
| Rural residence | 1323 (60·6) | 991 (65·5) | 332 (49·7) |
| Unemployed | 1437 (65·9) | 995 (65·7) | 442 (66·2) |
| Previously Prisoner<sup>#</sup> | 209 (9·6) | 90 (5·9) | 119 (17·8) |
| Education level |  |  |  |
| Primary | 770 (35·3) | 507 (33·5) | 263 (39·4) |
| Secondary | 1288 (59·0) | 912 (60·2) | 376 (56·3) |
| No. of household contacts (mean, SD) | 2·30 (2·33) | 2·47 (2·40) | 1·91 (2·11) |
| **Clinical** |  |  |  |
| Smear positive | 975 (44·7) | 697 (46·0) | 278 (41·6) |
| Drug resistance profiles<sup>§</sup> |  |  |  |
| Pan-susceptible | 1152 (52·8) | 939 (62·0) | 213 (31·9) |
| MDR | 779 (35·7) | 386 (25·5) | 393 (58·8) |

\* Fifty-four participants did not report the information of TB treatment history.
<sup>#</sup> One-hundred and eight participants did not report information of incarceration.
<sup>§</sup> The drug-resistant profiles in this table were determined by the whole-genome sequencing detection of the drug-resistant related mutations.

**Figure 1.**
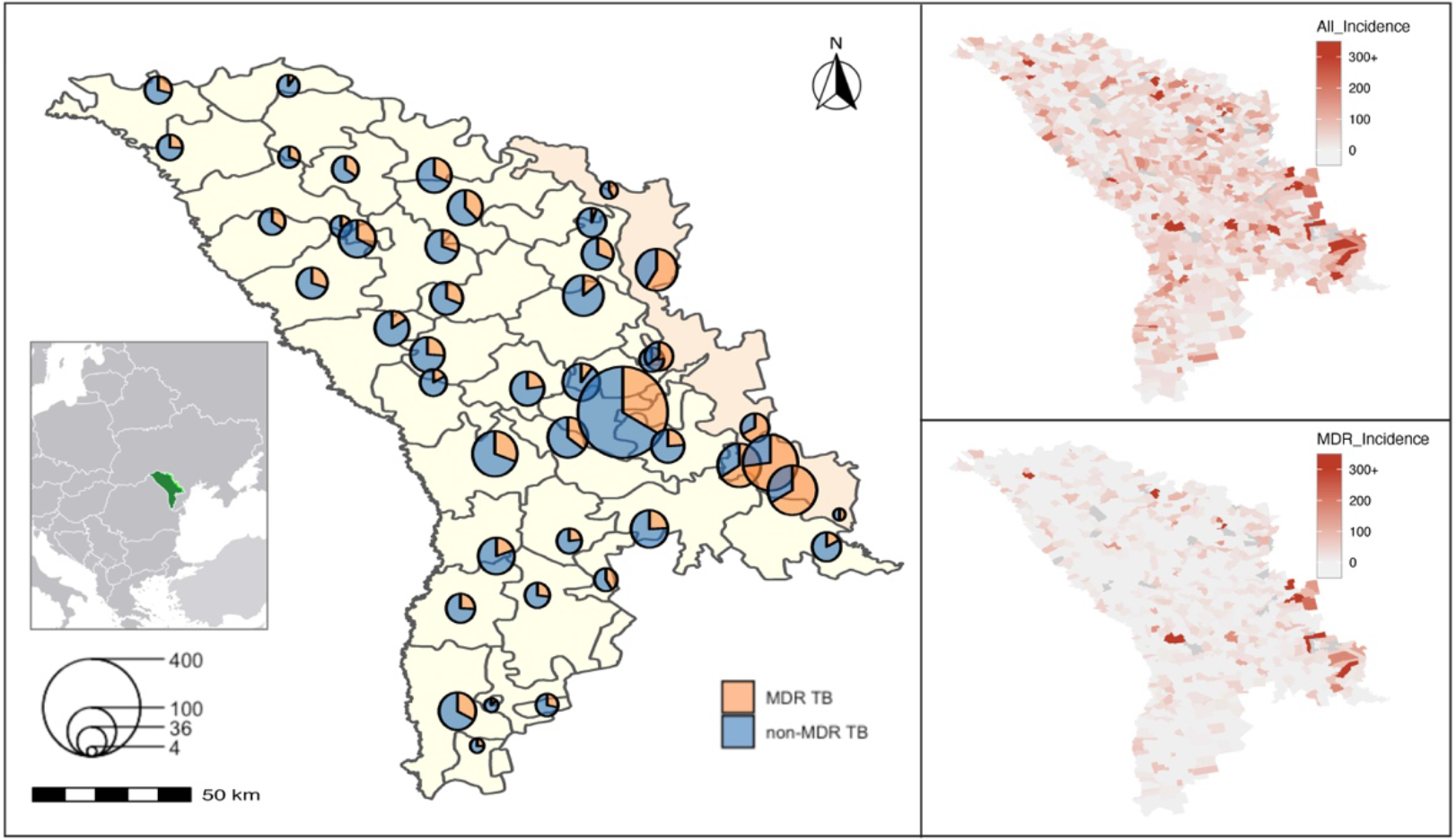
Map of culture-confirmed TB patients in Moldova (A). The center of each circle represents the geometric center of the localities/region where the case was diagnosed and sampled. The scale indicates the number of culture-confirmed TB patients. The Transnistrian region of Moldova is highlighted. The geographic distribution of the notified incidence of all culture-confirmed TB (B) and MDR-TB (C) by localities. The scheme of color represents the distribution of notified incidence and localities with missing population data are in gray.

779 participants (36%) were infected with genetic variants conferring MDR, 50% (386) of these MDR cases were treatment naÏve (**Table 1**). There was substantial geographic variation in distribution of MDR-TB. Transnistria, a small region east of the Dniester River, had localities with the highest proportions of TB cases that were MDR and among the highest incidence rates of MDR-TB in the country (**Fig. 1** and Supplementary **Fig. S1**).

### Genomic analysis and phylogeny reconstruction

We obtained sequence data from pre-treatment specimens of 2,220 participants. Polyclonal infections were identified in 386 participants (17·4%) (Supplementary Appendix, **Fig. S1** and **S2**) and removed, resulting in a final dataset of 1,834 *M. tuberculosis* isolates. Aligning reads against the reference strain revealed 43,284 SNPs that were used to reconstruct a M-L phylogeny (**Fig. 2**).

**Figure 2.**
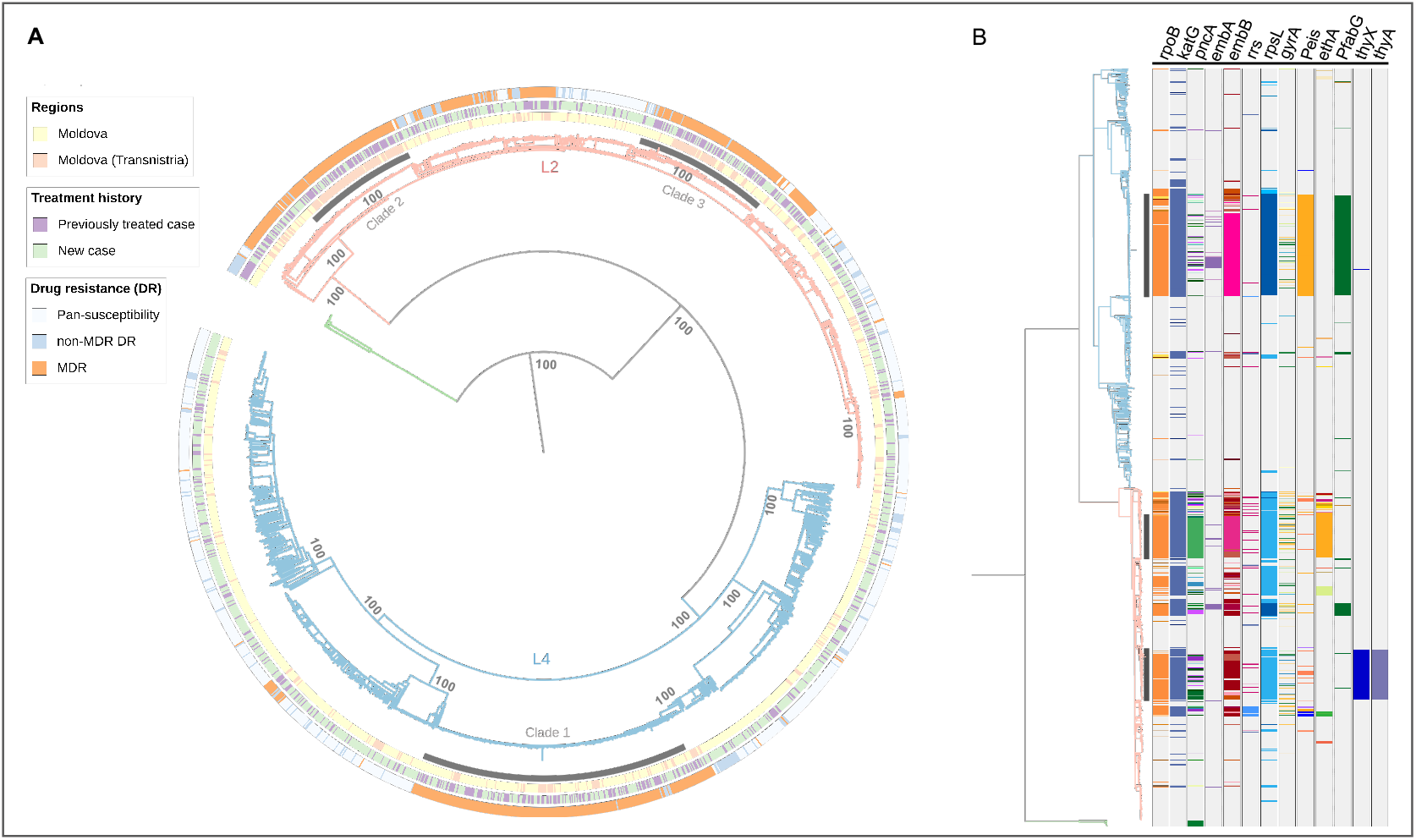
(A) Maximum-Likelihood (M-L) phylogeny of 1834 Moldova *M. tuberculosis* isolates based on 43,284 variable sites. The outer bands represent the *in-silico* drug-resistant profiles, treatment history of participant and the region where the isolates were sampled from. The tree is rooted to *M. bovis* (branch in green). L2 denotes lineage 2 (light orange) and L4 lineage 4 (light blue). Three major clades from the Ural/ lineage 4.2.1 (clade 1) and Beijing/lineage2.2.1 (clades 2-3) are shaded. The main nodes of the tree have 100% bootstrap support. (B) Phylogenetic distribution of resistance-related genotypes. The columns depict loci associated with drug resistance. “P” followed by a subscription of gene name indicates the promotor region. Colored bands of each column represent different polymorphisms.

1014 isolates (55·3%) belonged to Lineage 4 and 804 (43·8%) belonged to Lineage 2/sub-lineage 2.2.1 (**Fig. 2A**). Mapping revealed distinct geographic patterns for the three major MDR-TB clades: clade 1 comprising 243 Ural/lineage 4.2.1 isolates that were widely distributed, and clade 2 and clade 3 containing 102 and 121 Beijing/lineage strains that were concentrated within Transnistria (**Fig. 2A**). A high proportion of individuals (50.4%) in these three large MDR-TB clades had been previously treated for TB.

All Beijing/lineage 2.2.1 strains (802 consensus SNP call, 2 heterogenous SNP call) had a specific non-synonymous mutation in *esxW* (Thr2Ser), a gene in which mutations were found to be associated with transmission success of Beijing lineages in Vietnam.^21^ In contrast, just 3% of non-Beijing strains (32/1030) harbored this mutation (**Table S1**). Additionally, two non-synonymous variants in *esxW* were found in low frequencies in non-Beijing strains, six samples with a nonsense mutation at codon 172, and 17 samples with a Thr173Ser mutation.

### Prevalence of drug resistance genotypes

The three large clades were comprised almost entirely of MDR isolates (96%, 449 of 466) (**Fig.2B**); resistance-conferring mutations for isoniazid and rifampin were similar and found in the *katG* 315 codon and in the 81-bp rifampicin resistance-determination region (RRDR). However, each of these three clades had additional distinctive drug-resistance mutations: the isolates in Ural strain/lineage 4 clade 1 harbored an *eis* promoter (−12 C>T) mutation conferring kanamycin resistance, one Beijing strain/lineage 2 clade had an *ethA* (110-110 del), associated with ethionamide resistance, while the other had *thyX* (−16 C>T) and *thyA* (Arg222Gly) mutations, associated with resistance to p-aminosalicylic acid. We also identified clusters of isolates harboring additional drug-resistant mutations associated with drugs in newly-recommended MDR treatment regimens including lineozid (n=14), bedaquiline (n=1), and delamanid (n=9).

### Transmission of drug-resistant M. tuberculosis

Of the 1834 *M. tuberculosis* isolates, 1551 (85·6%) formed clusters ranging in size from two to 105, and 1000 (54·5%) belonged to 35 large clusters with at least ten participants at the clustering threshold of 0·001 substitutions/site. The median SNP distance across all transmission clusters was 14 SNPs (IQR 10-18 SNPs), with the median within-cluster SNP distance ranging from 0-26 SNPs (Supplementary Appendix, **Fig. S3**).

Of 672 MDR-TB isolates included in the final analysis, 619 (92·1%) were part of a cluster and 454 (67·6%) belonged to one of the 35 large transmission clusters. Individuals with MDR-TB were more likely to be in large clusters than individuals with pan-susceptible disease (Odds Ratio (OR) 3·39, *P*-value<0·01, Supplementary Appendix, **Table S2)**. Eight of the 14 MDR plus linezolid-resistant isolates were members of large clusters (Cluster 1, 2 and 21, Fig. S4). Among the nine MDR isolates with delamanid resistance, seven had the same delamanid-associated resistance mutation, forming a single sub-cluster (Cluster 19, **Fig. S4**) with a median pairwise SNP distance of < five SNPs, suggesting recent transmission of this highly-resistant *M. tuberculosis* strain in Moldova.

Closer inspection of the 35 large transmission clusters revealed distinct demographic and epidemiological differences between clusters. The largest transmission cluster (Cluster 1) included 105 participants with the sub-lineage 4.2.1/Ural Clade 1 stain residing throughout the entire country (**Fig. 3A and D**). In contrast, the next largest cluster (Cluster 2) included 102 participants with the sub-lineage 2.2.1/Beijing Clade 2 stain living predominately in Transnistria (**Fig. 3B and E**). Sixteen of the 35 large clusters were comprised almost entirely of MDR-TB (**Fig. 3 and S4**). Notably, there were cluster-specific demographic differences observed across transmission clusters, with the largest two groups comprising a high proportion of previous prisoners and reporting unsatisfactory living conditions (**Fig. S5**). **Table S2** details the association of covariates and membership in large clusters, along with a sensitivity analysis defining clusters using a stricter threshold of 0·0005 substitutions/site that showed broadly the same significant associations.

**Figure 3.**
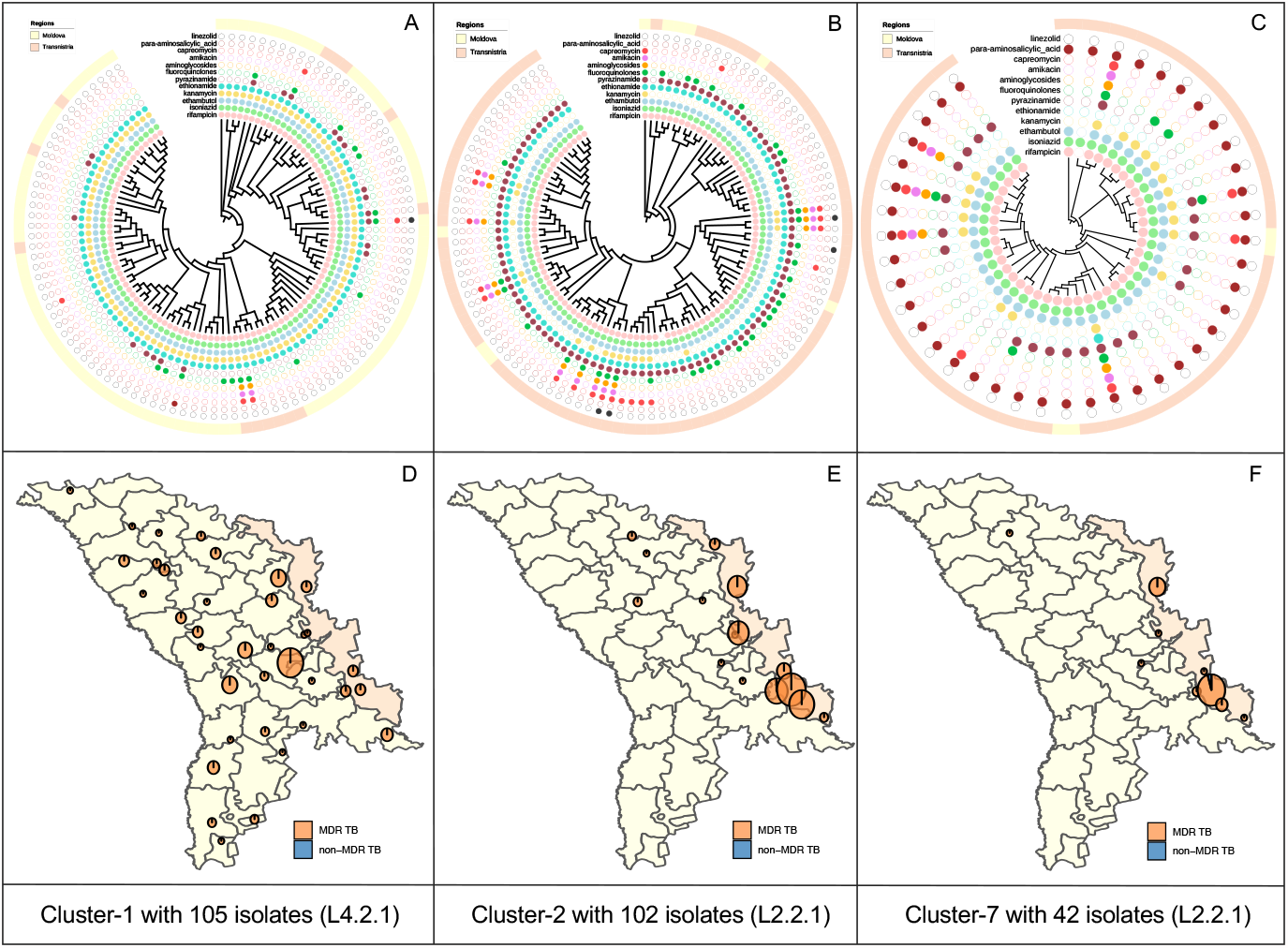
A-C: Tree visualizations for three large putative transmission clusters (N ≥ 10 isolates), each showing the location of cases in either the Moldova or Transnistria regions along with resistance/susceptibility to 12 anti-tuberculosis drugs, as identified by in silico prediction. D-E: Spatial distribution of three largest clusters (cluster 1, 2 and 7) in the Ural/Lineage 4.2.1 and Beijing/lineage 2.2.1 clades.

### Bayesian Skyline analysis of large MDR-TB clades

To gain further insight into the population dynamics of MDR-TB in Moldova, we reconstructed the scaled effective population size for the three large MDR-TB clades (**Fig. 2**) and estimated the time of the most recent common ancestor (TMRCA) (Supplementary Appendix, **Table S3**).

We estimated the TMRCA of the Ural/4.2.1 (clade 1) to be around 1984, though with a relatively broad posterior density interval (95% highest posterior density interval (HPDI)) of 1961 to 2003 owing to the limited temporal range of the data. The two Beijing/L2.2.1 clades (clades 2 and 3) are estimated to have a TMRCA of 2013 (95% HPDI: 2010-2015) and approximately 2006 (95% HPDI: 1999-2012), respectively (supplementary appendix, **Table S4**), implying more recent introduction of these clades to the region. **Fig. 4** shows the estimated *M. tuberculosis* effective population size for the three major clades over time. Our analysis estimated substantial growth of the Ural/clade 1 between 2012 and 2013 and of the Beijing/clade 3 in late-2013 to mid-2014. For the Beijing/clade 2, the effective population size has remained relatively constant, though the estimated date of origin falls within the time period when growth occurred in other clades. These results indicate a period of population expansion of MDR-TB in Moldova between 2012 to 2014. A sensitivity analysis using alternative clock models and rate estimates (Supplement Appendix **Table S4**) showed similar estimates for the TMRCA and effective population sizes for each clade (Supplementary Appendix **Fig. S6**).

**Figure 4:**
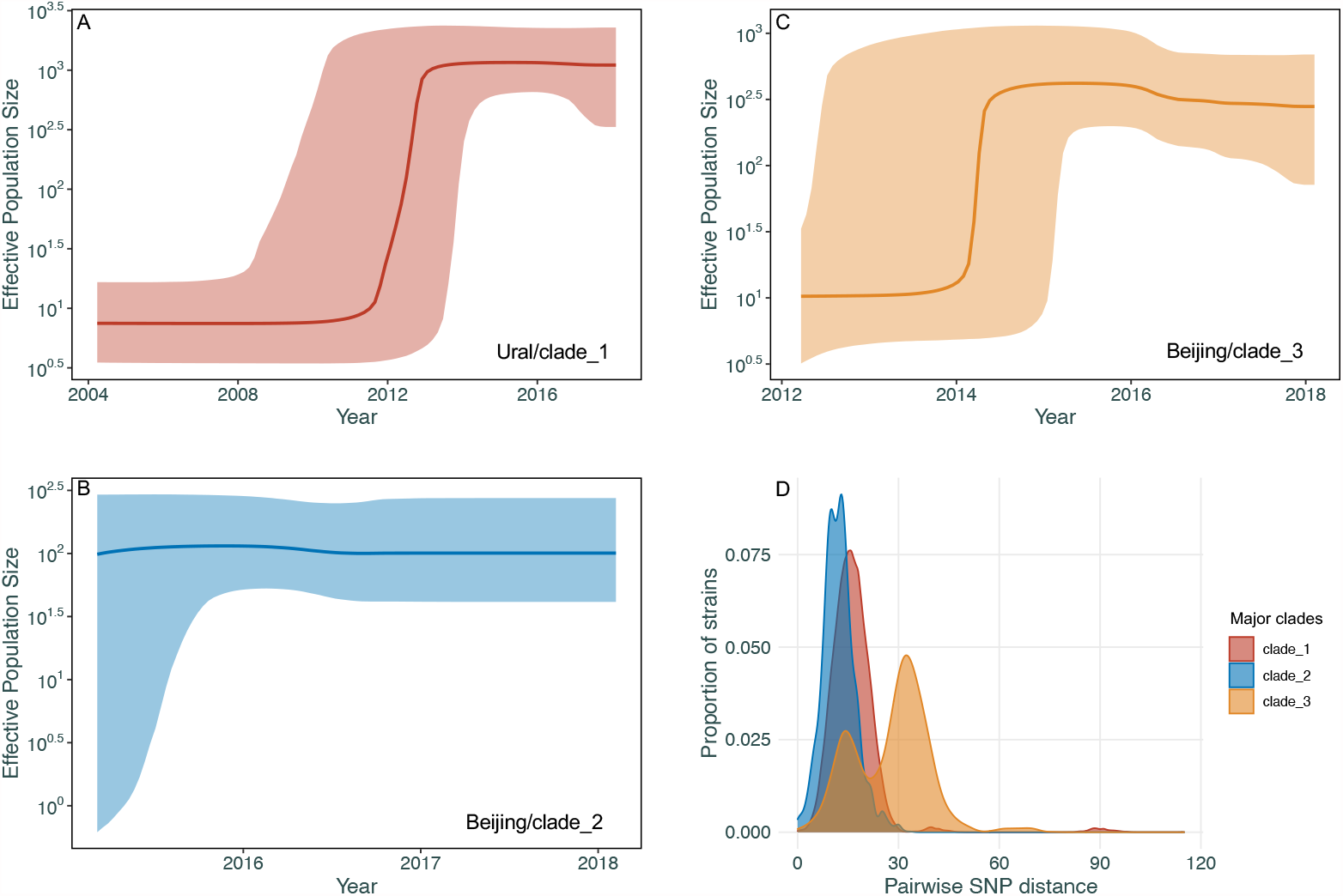
**A-C**: Coalescent Bayesian Skyline plots of the three large clades among Ural/lineage 4.2.1 and Beijing/lineage 2.2.1 with specific resistant mutations (detailed in **Fig. 2B**) using an uncorrelated log normal relaxed clock model. The two blue lines are the upper and lower bounds of the 95% HPD interval. The x-axis is the time in years and the y-axis is on a log-scale. **D**: Density distribution of within-clade isolates’ pairwise SNPs distance of Clade 1-3.

### Spatial/genetic distance analysis

Table 2 shows the pooled relative risk (RR) inference for pair- and individual-covariates from the Bayesian meta-analysis of genetic and spatial distances. Two cases in the same locality had a 46% lower expected patristic distance compared to cases in different localities (RR: 0·54; 95% CI: 0·39, 0·72). For cases in different localities, as the distance between the localities increases by 50 kilometers, the patristic distance between the pair increased by 7% (RR: 1·07 (1·03, 1·11)). For every half-year increase in the separation between dates of diagnosis for a pair, the patristic distance increased by 2% (RR: 1·02; 95% CI:1·01, 1·03). A sensitivity analyses using SNP distances yielded similar results (Supplementary Appendix, **Table S5**).

## Discussion

We describe the recent circulation of three distinct clades of *M. tuberculosis* (one of Ural lineage and two of Beijing lineage) responsible for the vast majority of MDR-TB in Moldova. While these clades share similar INH- and RIF-conferring mutations, there are additional clade-specific mutations conferring resistance to important second-line TB antibiotics critical for MDR treatment success.

**Table 2.** Pooled Bayesian meta-analysis inference for each effect on the relative risk scale. Posterior means and 95% quantile-based credible intervals are presented.

| Effect | Estimate | 95% Credible Interval |
| --- | --- | --- |
| Distance Between Localities (50 km) | 1·07 | (1·03, 1·11) |
| Same Locality (Yes vs. No) | 0·54 | (0·39, 0·72) |
| Date of Diagnosis Distance (1/2 year) | 1·02 | (1·01, 1·03) |
| Age Difference (10 years) | 1·01 | (0·99, 1·03) |
| Age (10 years) | 0·98 | (0·97, 1·00) |
| Household Contacts (1 person) | 0·99 | (0·97, 1·02) |
| Sex |  |  |
| Mixed Pair vs. Both Female | 0·96 | (0·90, 1·01) |
| Both Male vs. Both Female | 0·90 | (0·81, 0·99) |
| Residence Location |  |  |
| Mixed Pair vs. Both Not Urban | 1·06 | (0·99, 1·14) |
| Both Urban vs. Both Not Urban | 1·15 | (0·99, 1·32) |
| Housing |  |  |
| Mixed Pair vs. Both Not Homeless | 1·03 | (0·86, 1·25) |
| Both Homeless vs. Both Not Homeless | 1·15 | (0·84, 1·54) |
| Working Status |  |  |
| Mixed Pair vs. Both Unemployed | 1·04 | (0·95, 1·14) |
| Both Employed vs. Both Unemployed | 1·15 | (0·97, 1·36) |
| Education |  |  |
| Mixed Pair vs. Both ≥ Secondary | 0·97 | (0·93, 1·01) |
| Both $<$ Secondary vs. Both ≥ Secondary | 0·93 | (0·86, 1·02) |

Broad transmission networks based on genomic similarity showed that >85% of all culture-positive TB cases in Moldova could be mapped to putative transmission clusters, and that the majority (>54%) of these cases were found in 35 large transmission clusters. The role of recent transmission was even more pronounced for MDR-TB cases, among which >92% were found within putative transmission clusters (and >67% found within the 35 large transmission clusters). Individuals with MDR-TB had over three-fold higher odds of being in a large transmission cluster compared with individuals with pan-susceptible TB. Other notable covariates associated with increased odds of being in a large transmission cluster included urban residence, previous incarceration, and a history of previous treatment for TB. We found that pairs with closer times of diagnosis and living within the same locality had the greatest genomic similarity, and that for pairs in different localities, closer spatial proximity was associated with greater genomic similarity.

Previous analyses of surveillance data have revealed striking spatial heterogeneity of MDR-TB in Moldova with MDR-TB incidence differing by more than an order of magnitude for different localities,^22^ but the mechanisms driving this variation have not been described. Our analysis reveals that this heterogeneity is the result of multiple overlapping epidemics of transmitted MDR-TB, some of which are due to clades that have extended across the entire country, while others are thus far confined to specific subregions. Most notably, the two largest transmission clusters of the Beijing lineage are found almost exclusively in Transnistria, where, in some localities, MDR-TB incidence rates exceed 200 cases per 100,000 persons/year. Our finding that nearly all Beijing lineage strains in Moldova have *esxW* mutations corroborates recent work which suggests that these variants may be under positive selection.^21^

A recently-reported genomic study conducted among patients diagnosed in 2013 and 2014 at a single municipal hospital in Chisinau described the local concentration of MDR-enriched lineage 4.2.1 (Ural) isolates.^23^ In the current study, conducted approximately 6 years later and inclusive of the entire country, we found that MDR isolates within this lineage are present throughout Moldova and are commonly within transmission clusters, although this has thus far only been reported sporadically outside Moldova.^24^ Prior work had found this lineage to be responsible for MDR-TB due to reinfection in nosocomial settings;^25^ it is now apparent that these MDR strains are transmitted frequently in community settings. Regional reviews have suggested an important role of Beijing and Ural lineages in current TB epidemics; ^26^ our current work confirms and builds upon these insights, revealing in high resolution the overlapping dynamics of these two lineages in Moldova.

A major strength of our study was that we were able to include all culture-positive isolates across the country, minimizing challenges to transmission inference due to sampling biases. However, because we only could collect samples for two years—a short duration compared with the natural history of TB—our ability to track chains of transmission and to predict who infected whom was limited.

There are urgent clinical and public health implications of these findings. While the crisis of transmitted MDR-TB was already apparent in this region, these data reveal that there are several co-circulating highly-drug resistant TB clades that differ in terms of drug-resistance profiles, geographic distribution, and epidemic trajectory. These results suggest the urgency of interrupting MDR-TB transmission in Moldova, especially within specific geographic foci in the capital city of Chisinau and in the region of Transnistria. While the role of genomic surveillance for informing TB interventions in high-burden settings remains incompletely explored, this study provides an important example of how such information may be used to understand the complex epidemiology of MDR-TB in a high incidence country.

## Data Availability

The sequencing data will be made available through GenBank. Additional data can be requested from the corresponding authors who reserve the right to decide whether to share data based on the materials provided by researchers.

## Data sharing

The sequencing data will be made available through GenBank (PRJNA736718). Additional data can be requested from the corresponding authors who reserve the right to decide whether to share data based on the materials provided by researchers.

## Acknowledgements

We thank the clinical and laboratory staff of Phthisiopneumology Institute from Chisinau and Regional Reference Laboratories from Balti, Vorniceni and Bender from Moldova for invaluable help and for their assistance in collecting and testing patient specimens. This study was made possible by the generous support of the American people through the United States Agency for International Development (USAID) through the TREAT TB Cooperative Agreement No. GHN-A-00-08-00004. The contents are the responsibility of the authors and Subgrantee and do not necessarily reflect the views of USAID or the United States Government.

