## Supplemental for "Phylogeography and transmission of *M. tuberculosis* in Moldova"

**Supplementary appendix**

**S1. Study population**

All individuals evaluated for tuberculosis, with the exception of those in detention facilities, in the Republic of Moldova in 2018 and 2019 were invited to participate in the study. Consenting individuals with positive sputum cultures were included in this study.

**Fig. S1.** Distribution of the proportion of MDR-TB by the regions where they were diagnosed. (A) Regions sorted by the proportion of MDR-TB and (B) the total numbers of MDR-TB isolates from high to low.

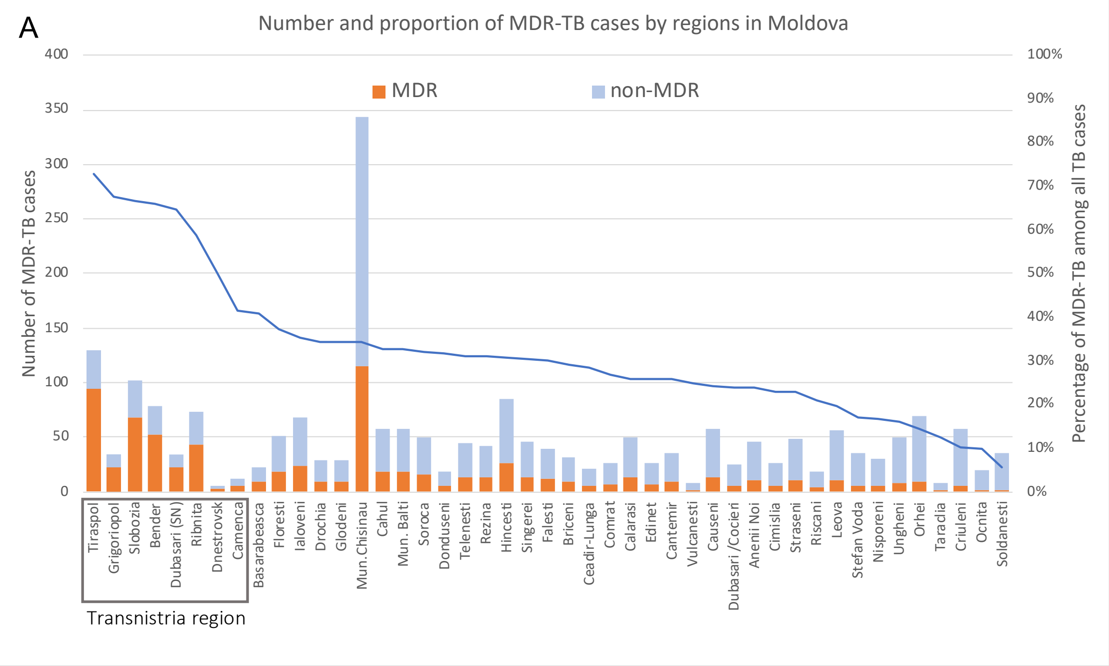


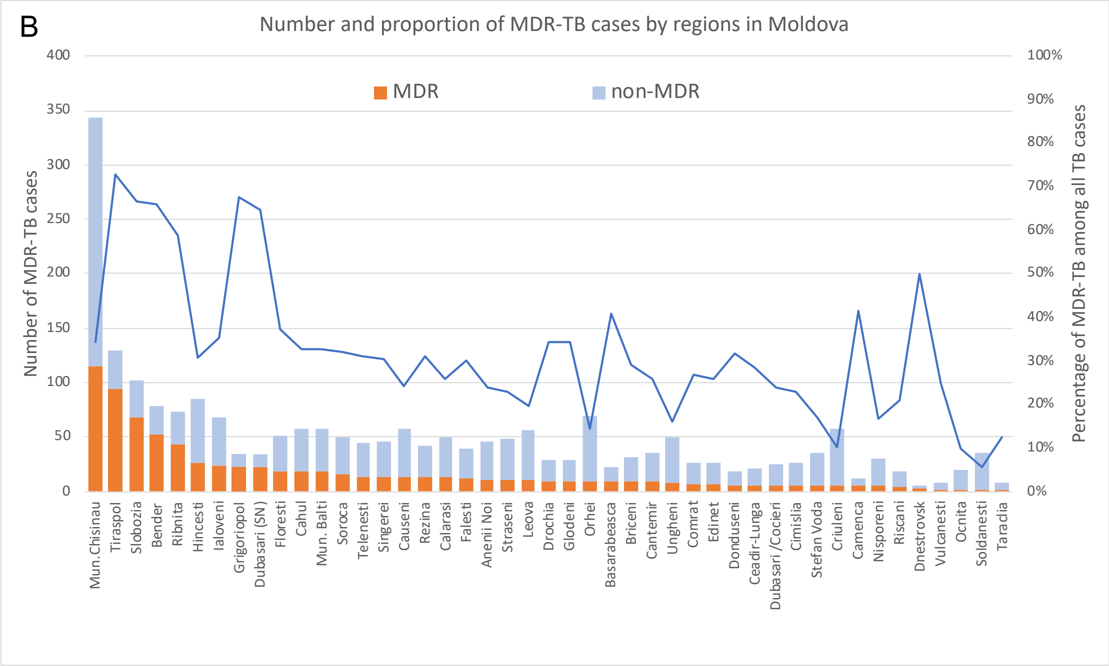

### S2. Whole-genome sequencing analysis

*Whole-genome DNA preparation methods*

Genomic DNA was prepared for whole genome sequencing using the Illumina DNA Prep library preparation kit. Libraries were indexed using the Illumina DNA Prep Unique Dual Indexing system, pooled at equimolar concentrations, and sequenced on an Illumina NextSeq 500 instrument. Up to 285 pooled samples were sequenced on a single NextSeq High Output 300 cycle sequencing kit.

To decrease cost per sample, the Illumina DNA Prep protocol was modified to reduce reagent usage to one fourth of the recommended volumes. Validation of this method showed lower concentration of the final libraries, however no significant loss in genome quality (coverage depth, coverage breadth, sequence duplication levels) was observed compared to full reaction volumes.

*Identifying putative mixed infection*

Preliminary results from an initial genomic analysis of all samples with whole genome sequence data (N = 2236) showed a discordance between the observed pairwise genomic variation (SNP distance) between isolates and their patristic distance on a well-supported maximum-likelihood phylogeny (Figure S2-A). We aimed to improve the relationship between genomic variation and phylogenetic relatedness by identifying and removing putative mixed infection in our sampled population. We employed the approach detailed in Sobkowiak *et. al.* 2018 for detecting the signal of mixed infection from whole genome sequence data.

Briefly, this method computed the likelihood of a sample containing two or more distinct *Mtb* strains by extracting allele frequencies from the sequencing reads at each position that has been called as a heterogenous site ('0/1') from the GATK variant calling pipeline. A Bayesian clustering approach is then applied to these per-sample heterogenous allele frequencies to determine the likelihood of these frequencies clustering into two or more distinct groups with a mean frequency between 0.2 and 0.8 (indicative of polyclonal infection), or one highly dispersed group (likely clonal variation). The results of this analysis identified 403 possible mixed infections within our sampled population (18.0%), resulting in 1834 non-mixed isolates that were included for the main analysis that showed a strong concordance in the pairwise SNP distance and patristic distance in a maximum-likelihood phylogeny (Figure S2-B).

*BEAST analysis*

Large putative clusters (≥ ten cases) obtained using TreeCluster [18] from a patristic distance threshold of 0.001 substitutions/site were individually analyzed to build timed phylogenetic trees with BEAST2 v2.6.3.[19]. All sequences passed a test for homogeneity of nucleotide composition using IQ-TREE v1.6.12,[20]. Phylogenies were built using a strict molecular clock, calibrated by the tip date of collection, and a fixed clock rate parameter of $1.0\times10^{-7}$ per site per year, equating to 0.44 substitutions per genome per year [21] across all the clusters. We used a coalescent constant population model with a log normal [0,200] prior distribution[21] and a correction for ascertainment bias. We ran the Markov chain Monte Carlo (MCMC) algorithm for 250 million iterations and retained every 25,000-th steps from the posterior. A maximum clade credibility (MCC) tree was generated with the help of TreeAnnotator v2.6.2,[19] with 10% of the chain discarded as burn-in. We used a coalescent Bayesian Skyline model to infer the events of *M. tuberculosis* population expansion to estimate the effective population size change through time in three large clades that were identified in the study population that contained individuals with specific drug resistance mutations. The same strategy was adopted for checking sequence homogeneity composition and optimal model selection (Table S3), and each clade was analyzed separately.

Figure S2-A. A scatterplot showing the pairwise SNP distance (max. 50 SNP differences) plotted against the patristic distance on a maximum-likelihood phylogeny produced with RAxML between all 2237 Moldovan isolates with whole genome sequence data.

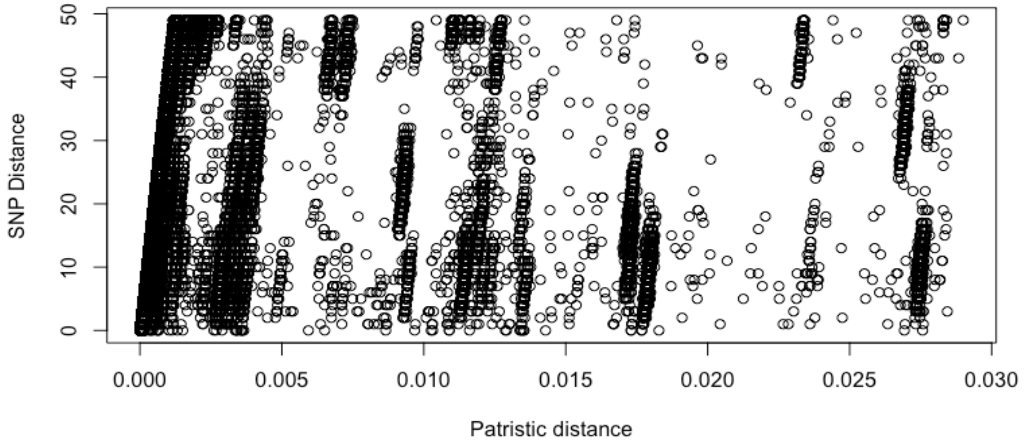


Figure S2-B. A scatterplot showing the pairwise SNP distance (max. 50 SNP differences) plotted against the patristic distance on a maximum-likelihood phylogeny produced with RAxML between 1834 non-mixed Moldovan isolates with whole genome sequence data.

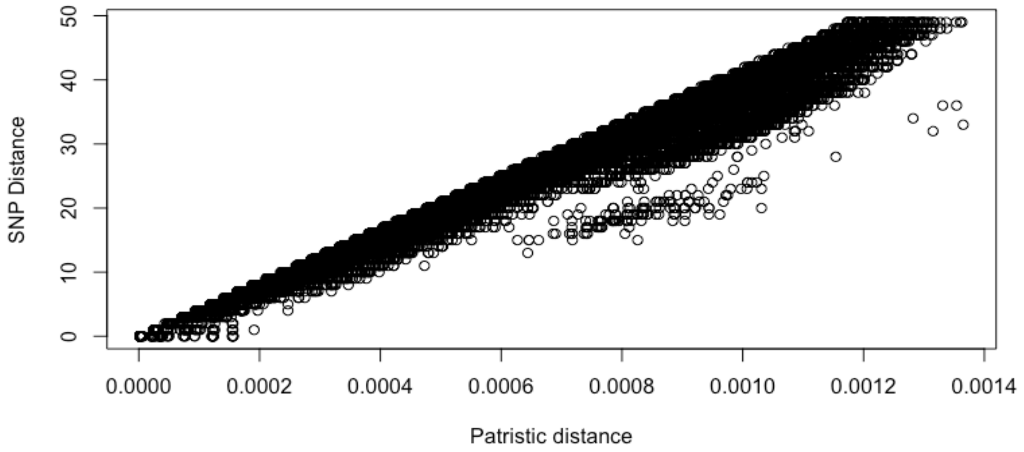

**S2. Genomic clustering analysis**

Fig. S3. The pairwise SNP distance in 35 large transmission clusters with at least 10 participants involved. The box plot shows the IQR and median SNP distance of each cluster.

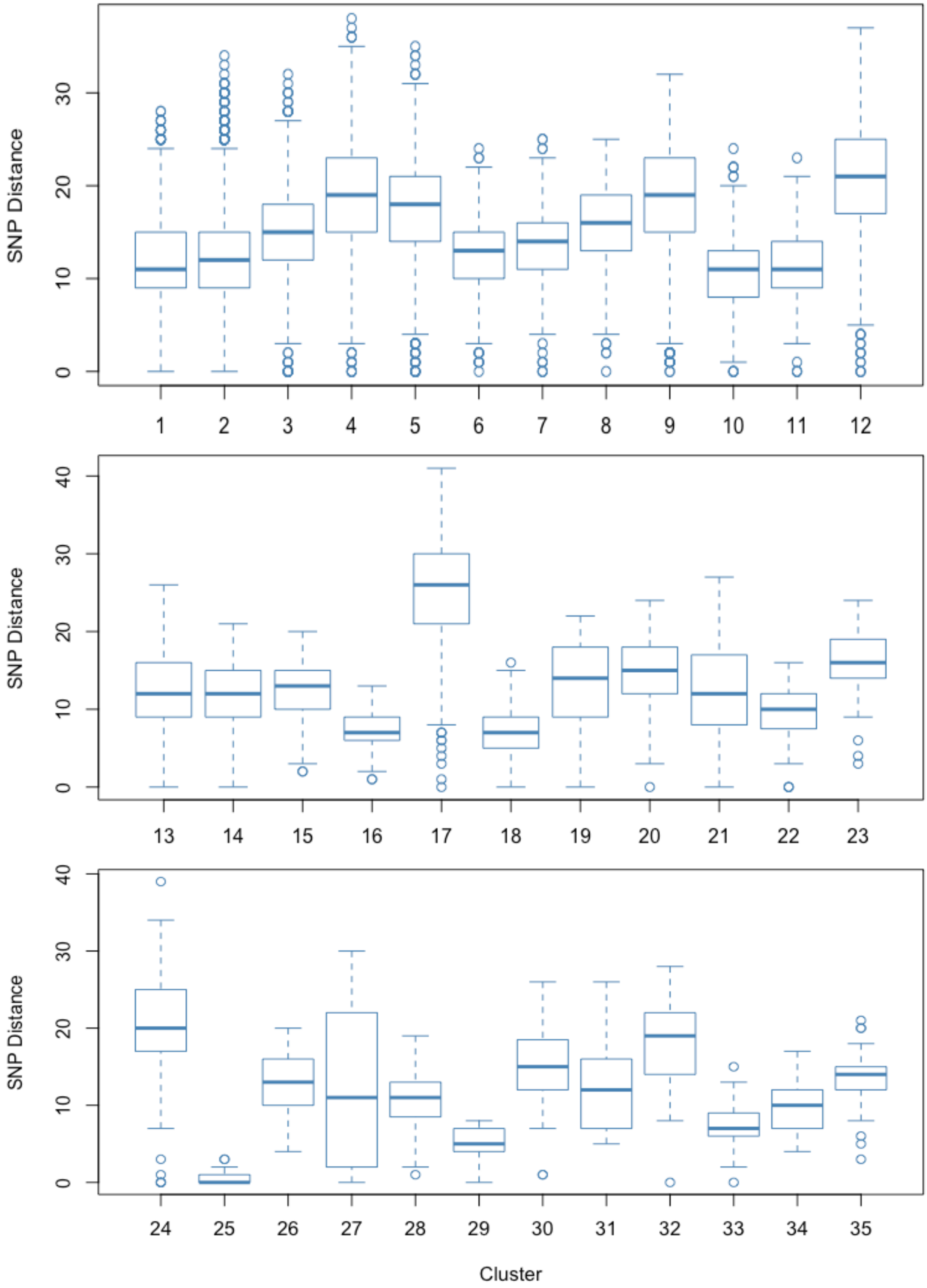

**Table S1.** Allele counts for nine SNP variants identified in the *esxW* gene within the study population, showing counts within samples classified as either Beijing strains (all lineage 2.2.1) or as any other lineage.

| | | | Beijing strain (N = 804) | | | | non-Beijing strains (N = 1030) | | | |
|---|---|---|---|---|---|---|---|---|---|---|
| **H37Rv Index** | **Mutation** | **Syn/non-syn** | **Reference allele count** | **Alternative allele count** | **Mixed call** | **Missing call** | **Reference allele count** | **Alternative allele count** | **Mixed call** | **Missing call** |
| 4060334 | T-C | S | 803 | 0 | 0 | 1 | 803 | 213 | 0 | 14 |
| 4060417 | G-A | NS | 804 | 0 | 0 | 0 | 1012 | 6 | 0 | 12 |
| 4060418 | G-T | S | 804 | 0 | 0 | 0 | 1017 | 1 | 0 | 12 |
| 4060420 | T-C | NS | 804 | 0 | 0 | 0 | 1001 | 17 | 0 | 12 |
| 4060466 | G-A | S | 804 | 0 | 0 | 0 | 990 | 24 | 4 | 12 |
| 4060469 | G-C | S | 804 | 0 | 0 | 0 | 1016 | 1 | 1 | 12 |
| 4060472 | A-G | S | 804 | 0 | 0 | 0 | 1016 | 1 | 1 | 12 |
| 4060545 | G-T | S | 804 | 0 | 0 | 0 | 1015 | 1 | 0 | 14 |
| 4060562 | G-A | S | 804 | 0 | 0 | 0 | 1010 | 5 | 0 | 15 |
| 4060588 | T-C | NS | 0 | 802 | 2 | 0 | 984 | 31 | 1 | 14 |

**Table S2** detailed the associated characteristics with the large clusters. Membership in large clusters was found to be significantly associated with age group, previous history of TB treatment and previous imprisonment. Individuals between the ages of 20-29 and 30-29 were more likely to be in large clusters (OR 2·64 and 2·21, *P* value <0·01), as were individuals that either had TB diagnosed as a relapse or treatment failure compared to new cases (OR 1·85 and 2·54, *P* value <0·01), and those that had a history of imprisonment than those that did not (OR 2·57, *P* value <0·01). In addition, individuals in large clusters were more likely to live in Transnistria than the rest of Moldova (OR 3·57, *P* value <0·01) and to reside in an urban center (OR 1·57, *P* value <0·01).

**Table S2**. Demographic associations in cases belonging to large transmission clusters (≥ 10 cases), identified with patristic distance thresholds of 0.001 and 0.0005. Cases in small clusters (2 – 9 cases) are not included. Odds ratios are calculated using logistic regression and *P* values by Wald chi-squared test, adjusted for age and sex.

| | | **No. in large cluster (cutoff 0·001)** | **Odds ratio** | **P value** | **No. in large cluster (cutoff 0·0005)** | **Odds ratio** | **P value** |
|---|---|---|---|---|---|---|---|
| **Total** | | 1000/1283 | | | 404/951 | | |
| **Age** | | | | | | | |
| | <20 | 24/31 | 1·60 (0·70 - 4·16) | | 4/17 | 0·78 (0·22 - 2·33) | |
| | 20-29 | 129/151 | 2·64 (1·62 - 4·47) | | 56/102 | 2·94 (1·84 - 4·70) | |
| | 30-39 | 313/375 | 2·21 (1·56 - 3·14) | | 129/259 | 2·30 (1·63 - 3·27) | |
| | 40-49 | 242/314 | 1·45 (1·03 - 2·05) | | 111/255 | 1·75 (1·23 - 2·49) | |
| | 50+ | 268/384 | 1 | **<0·01** | 88/291 | 1 | **<0·01** |
| **Sex** | | | | | | | |
| | Male | 768/980 | 1 | | 318/732 | 1 | |
| | Female | 232/303 | 0·79 (0·58 - 1·09) | 0·15 | 86/219 | 0·70 (0·50 - 0·97) | **0·03** |
| **TB type** | | | | | | | |
| | New case | 637/856 | 1 | | 249/641 | 1 | |
| | Relapse | 262/312 | 1·85 (1·32 - 2·63) | | 99/214 | 1·38 (1·00 - 1·90) | |
| | Treatment failure | 76/86 | 2·54 (1·35 - 5·32) | **<0·01** | 39/68 | 2·09 (1·26 - 3·51) | **<0·01** |

| | | | | | | | |
|---|---|---|---|---|---|---|---|
| **Smear status** | | | | | | | |
| | Positive | 412/540 | 0·96 (0·76 - 1·28) | | 169/409 | 1·19 (0·90 - 1·59) | |
| | Negative | 440/570 | 1 | 0·8 | 159/420 | 1 | 0·23 |
| **Drug resistance** | | | | | | | |
| | Sensitive | 456/648 | 1 | | 166/476 | 1 | |
| | Drug-resistant | 90/129 | 0·83 (0·55 - 1·29) | | 43/110 | 0·86 (0·53 - 1·38) | |
| | MDR | 454/506 | 3·39 (2·44 - 4·79) | <0·01 | 195/365 | 2·03 (1·53 - 2·70) | **<0·01** |
| **Location** | | | | | | | |
| | Moldova | 713/967 | 1 | | 293/757 | 1 | |
| | Transnistria | 287/316 | 3·57 (2·40 - 5·49) | **<0·01** | 111/194 | 2·14 (1·55 - 2·97) | **<0·01** |

Table S2. continued

| | | No. in large cluster (0·001) | Odds ratio | P value | No. in large cluster (0·0005) | Odds ratio | P value |
|---|---|---|---|---|---|---|---|
| **Urban dwelling** | | | | | | | |
| | Yes | 427/517 | 1·57 (1·18 - 2·10) | | 174/378 | 1·30 (0·99 - 1·70) | |
| | No | 549/736 | 1 | **<0·01** | 214/544 | 1 | 0·06 |
| **Homeless** | | | | | | | |
| | Yes | 97/119 | 1·25 (0·78 - 2·08) | | 39/92 | 0·95 (0·61 - 1·48) | |
| | No | 857/1104 | 1 | 0·37 | 342/811 | 1 | 0·83 |
| **Monetary assistance** | | | | | | | |
| | Yes | 284/372 | 1·08 (0·79 - 1·48) | | 110/276 | 1·06 (0·78 - 1·44) | |
| | No | 613/780 | 1 | 0·62 | 252/580 | 1 | 0·72 |
| **Living conditions** | | | | | | | |
| | Satisfactory | 459/583 | 1·05 (0·79 - 1·41) | | 169/423 | 0·81 (0·60 - 1·07) | |
| | Unsatisfactory | 414/532 | 1 | 0·74 | 173/382 | 1 | 0·14 |
| **Occupation** | | | | | | | |
| | Employed | 121/155 | 1 | | 53/115 | 1 | |
| | Disabled | 95/120 | 1·16 (0·65 - 2·10) | | 35/88 | 0·81 (0·46 - 1·43) | |
| | Retired | 79/123 | 0·73 (0·41 - 1·27) | | 26/98 | 0·63 (0·34 - 1·16) | |
| | Student | 22/27 | 0·65 (0·23 - 2·16) | | 3/13 | 0·19 (0·04 - 0·68) | |
| | Unemployed | 655/825 | 1·02 (0·66 - 1·54) | 0·44 | 269/606 | 0·86 (0·57 - 1·29) | 0·11 |
| **Education** | | | | | | | |
| | Primary | 338/441 | 1 | | 134/317 | 1 | |
| | Secondary | 411/537 | 1·01 (0·74 - 1·36) | | 154/399 | 0·86 (0·63 - 1·16) | |
| | Specialized secondary | 169/204 | 1·52 (1·00 - 2·36) | | 73/153 | 1·26 (0·85 - 1·87) | |
| | Higher education | 34/42 | 1·23 (0·57 - 2·95) | | 11/27 | 0·91 (0·39 - 2·03) | |
| | No education | 19/20 | 5·18 (1·04 - 93·98) | 0·15 | 12/16 | 3·83 (1·28 - 14·08) | 0·04 |
| **Previously prisoner** | | | | | | | |
| | Yes | 114/127 | 2·57 (1·46 - 4·90) | | 48/84 | 1·87 (1·18 - 2·99) | |
| | No | 772/1018 | 1 | **<0·01** | 302/757 | 1 | **<0·01** |

**Table S3.** Results of the Coalescent Bayesian Skyline analyses of the three large clades with specific resistant mutations using an uncorrelated log normal relaxed clock model.

| Clades based on DR mutations | # of Taxa | Substitution Model | tMRCA | Clock Rate (SNPs per site per year) |
|---|---|---|---|---|
| Ural Clade 1 | 243 | TIM with unequal base frequencies | mean: 1984, 95% HPD interval: 1961 - 2003 | mean: $2.805 \times 10^{-7}$, 95% HPD interval: $1.636 \times 10^{-7}$ - $3.954 \times 10^{-7}$ |
| Beijing Clade 2 | 102 | TVM with equal base frequencies | mean: 2013, 95% HPD interval: 2010 - 2015 | mean: $6.248 \times 10^{-7}$, 95% HPD interval: $2.445 \times 10^{-7}$ - $1.087 \times 10^{-6}$ |
| Beijing Clade 3 | 121 | TVM with equal base frequencies | mean: 2006, 95% HPD interval: 1999 - 2012 | mean: $5.005 \times 10^{-7}$, 95% HPD interval: $2.658 \times 10^{-7}$ - $7.594 \times 10^{-7}$ |

### S3. Phylogenetic reconstruction and Bayesian Skyline analysis

**Fig. S4:** Tree visualizations for remaining 32 transmission clusters (N ≥ 10 isolates), each showing the location of cases in either the Moldova or Transnistria regions along with resistance/susceptibility to anti-tuberculosis drugs, as identified by *in silico* prediction.

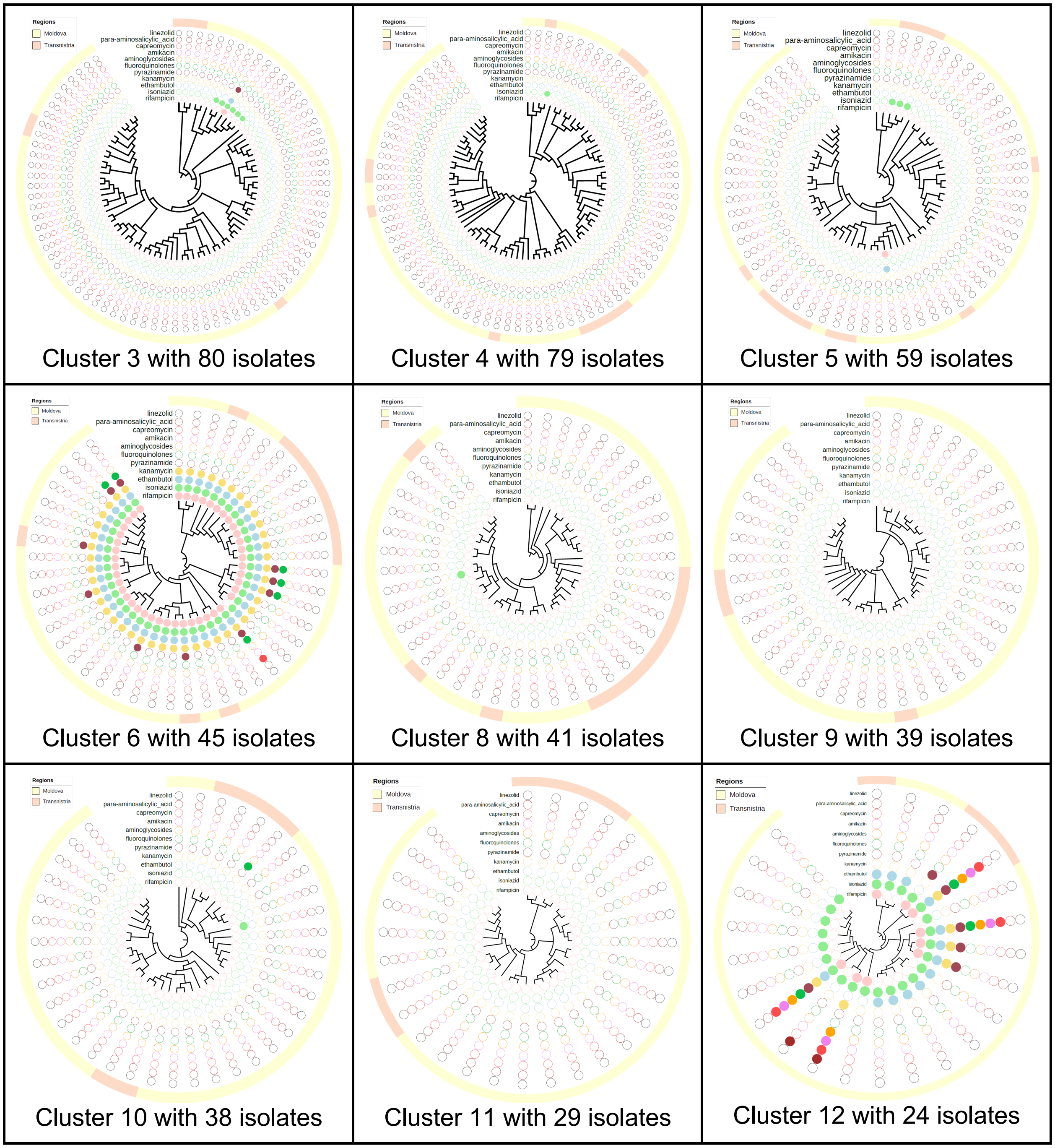

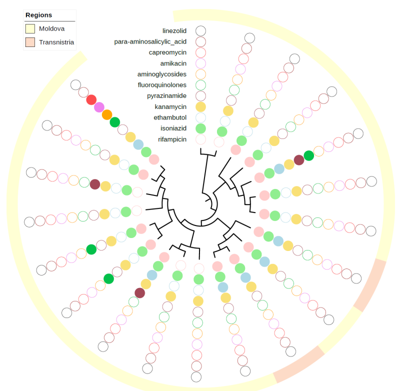

Cluster 13 with 20 isolates

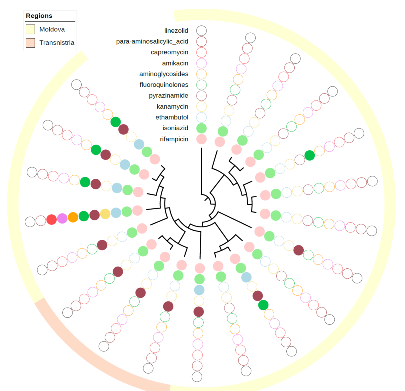

Cluster 14 with 20 isolates

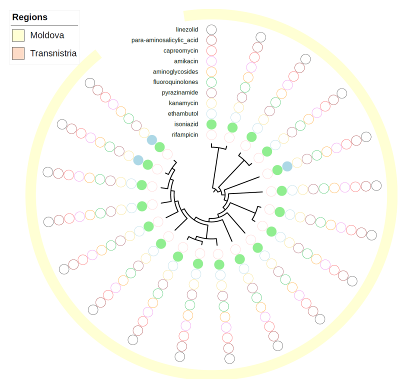

Cluster 15 with 19 isolates

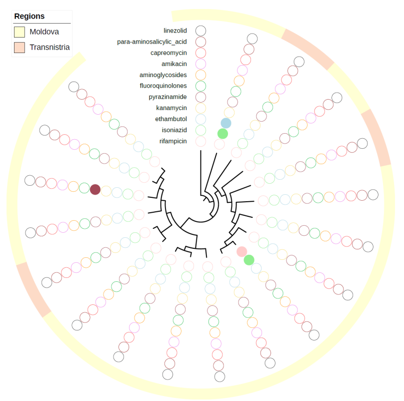

Cluster 16 with 19 isolates

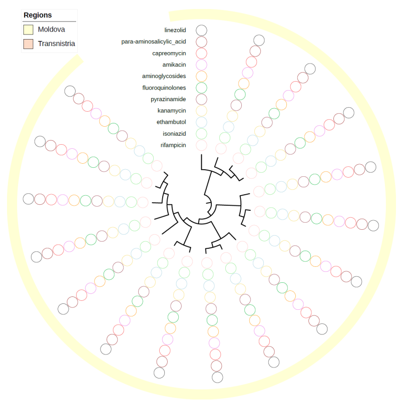

Cluster 17 with 17 isolates

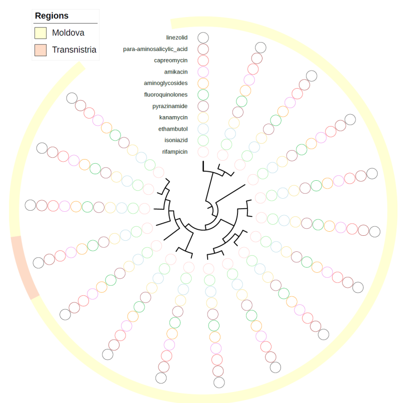

Cluster 18 with 17 isolates

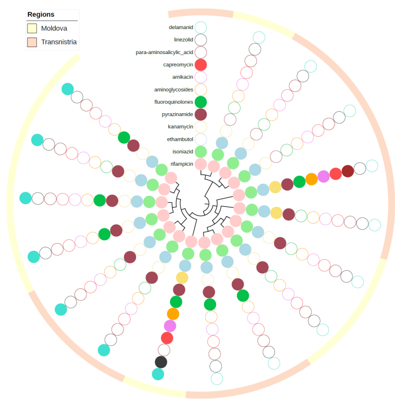

Cluster 19 with 17 isolates

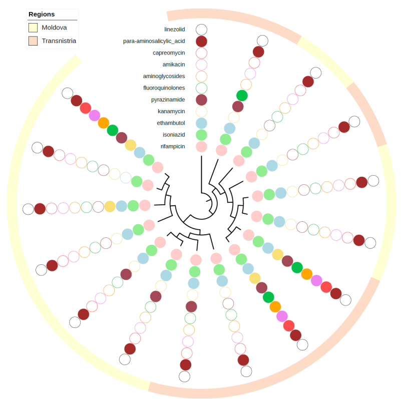

Cluster 20 with 16 isolates

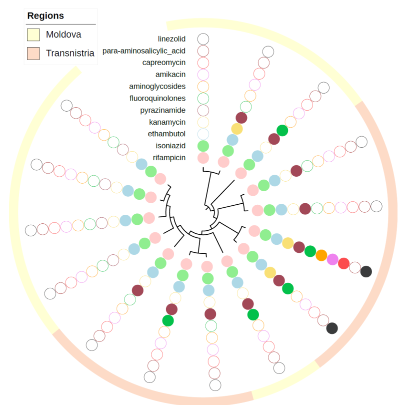

Cluster 21 with 15 isolates

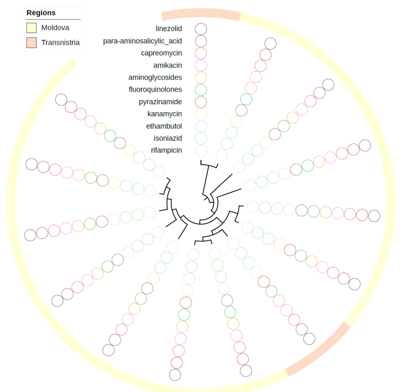


Cluster 22 with 14 isolates

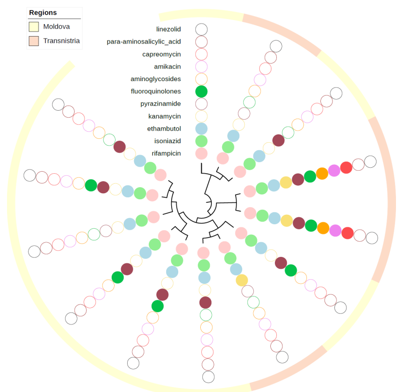


Cluster 23 with 13 isolates

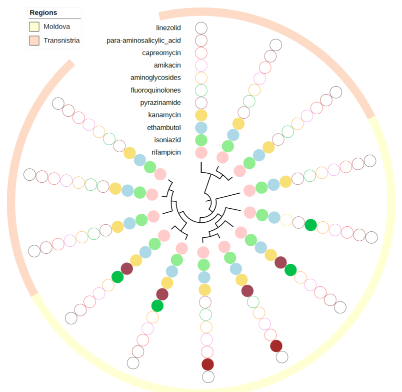


Cluster 24 with 13 isolates

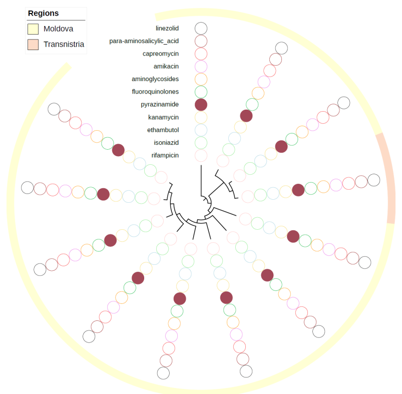


Cluster 25 with 12 isolates

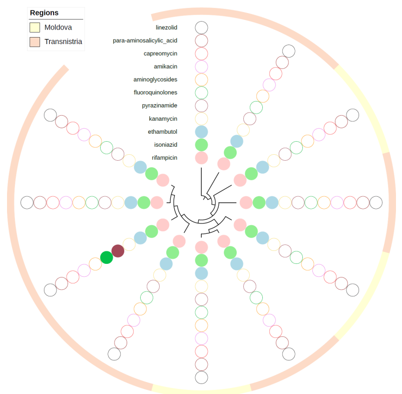


Cluster 26 with 11 isolates

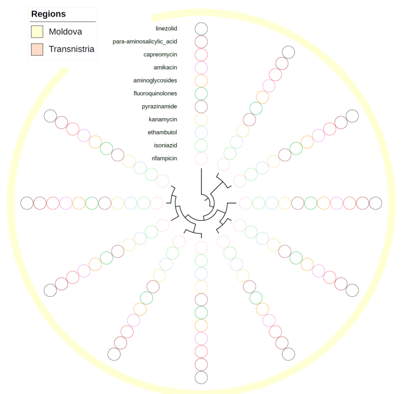


Cluster 27 with 11 isolates

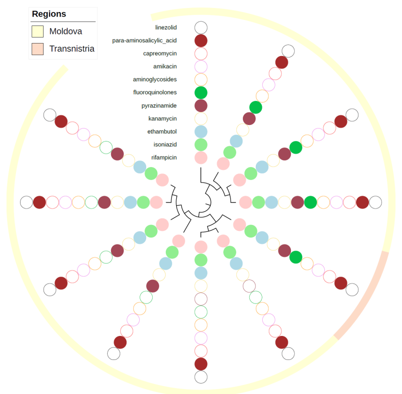


Cluster 28 with 11 isolates

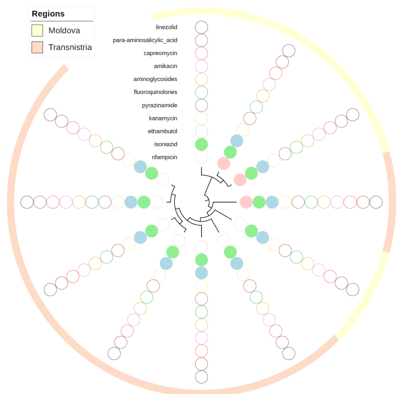


Cluster 29 with 11 isolates

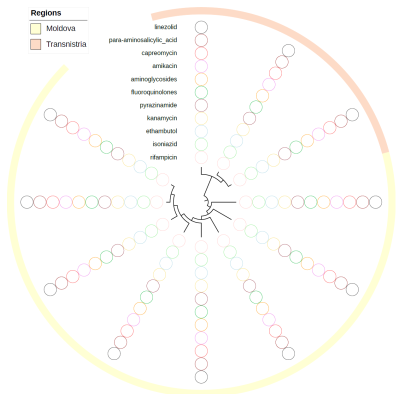


Cluster 30 with 11 isolates

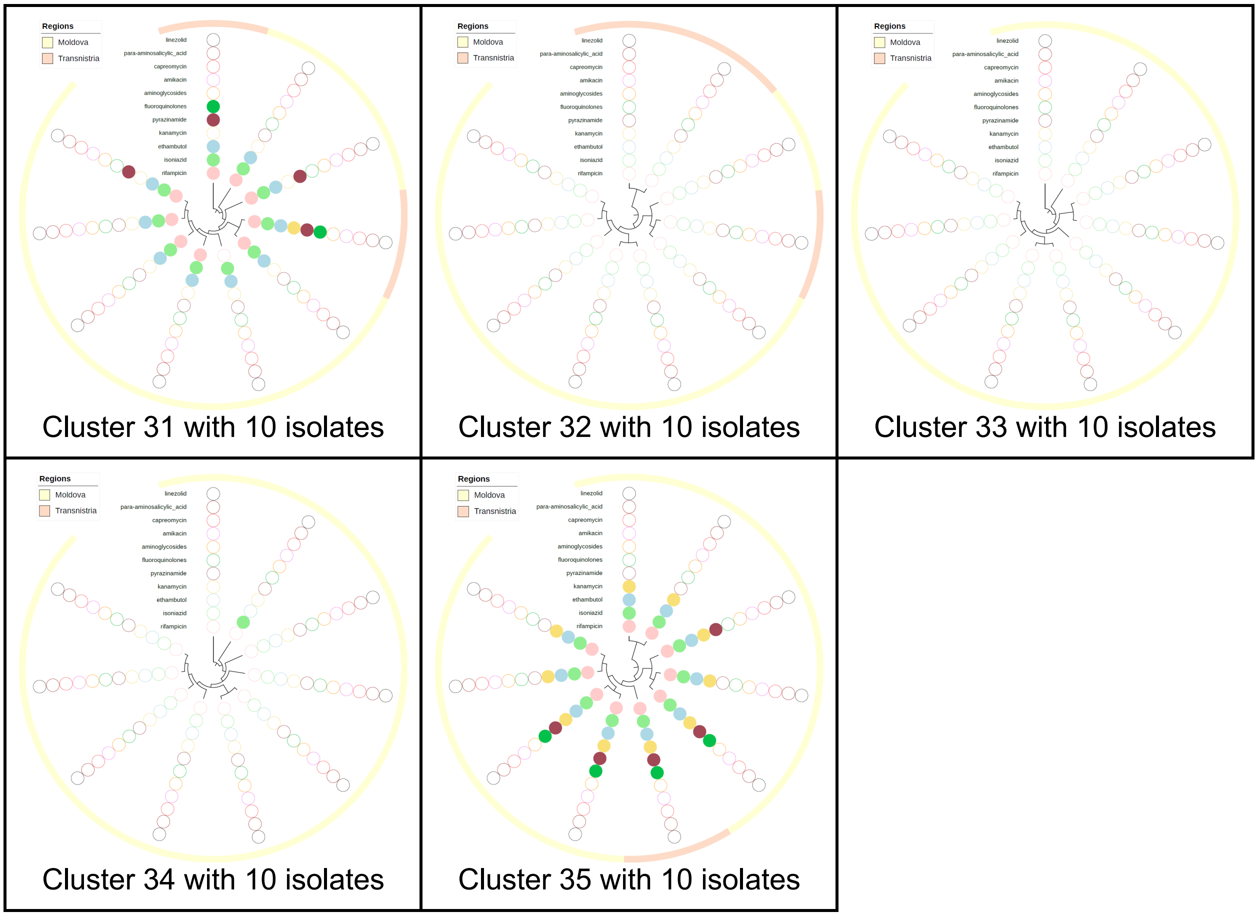
Regions
Moldova
Transnistria
linezolid
para-aminosalicylic_acid
capreomycin
amikacin
aminoglycosides
fluoroquinolones
pyrazinamide
kanamycin
ethambutol
isoniazid
rifampicin
Cluster 31 with 10 isolates
Cluster 32 with 10 isolates
Cluster 33 with 10 isolates
Cluster 34 with 10 isolates
Cluster 35 with 10 isolates

**Fig. S5**: Tree visualizations for the 35 transmission clusters (N ≥ 10 isolates), each showing the location of cases in either the Moldova and Transnistria regions along with selected covariates, namely, urban residence, homeless, unsatisfactory living conditions and former prisoner.

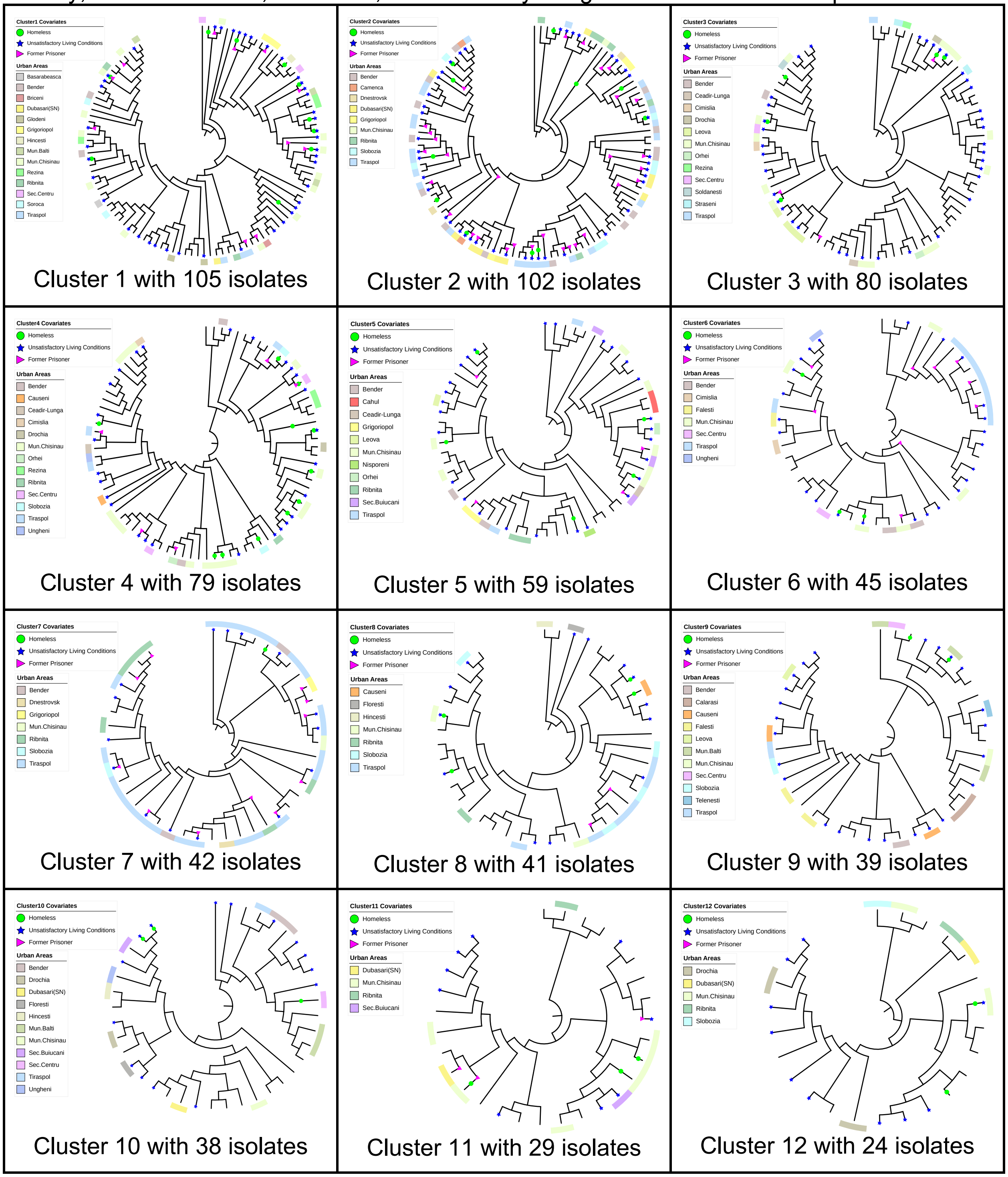

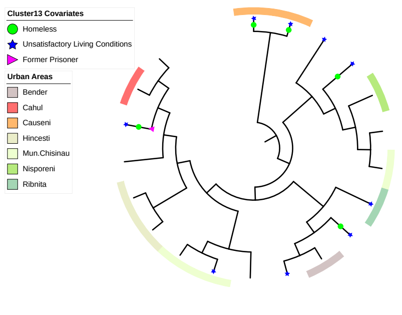


Cluster 13 with 20 isolates

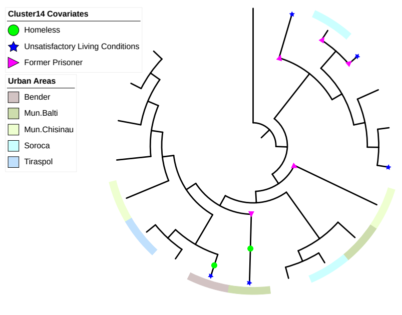


Cluster 14 with 20 isolates

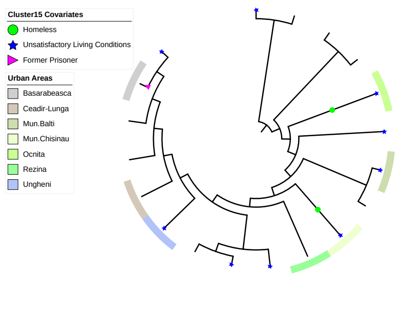


Cluster 15 with 19 isolates

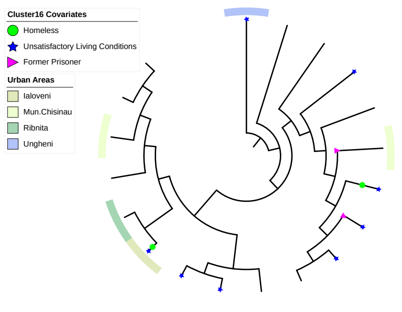


Cluster 16 with 19 isolates

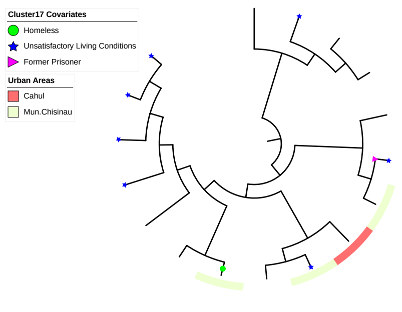


Cluster 17 with 17 isolates

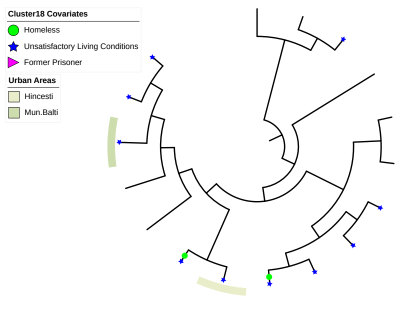


Cluster 18 with 17 isolates

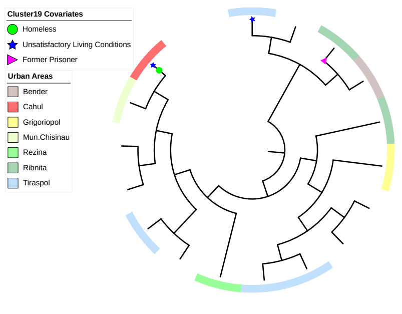


Cluster 19 with 17 isolates

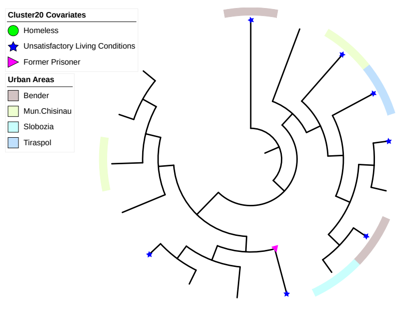


Cluster 20 with 16 isolates

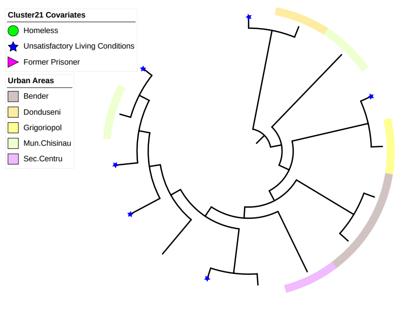


Cluster 21 with 15 isolates

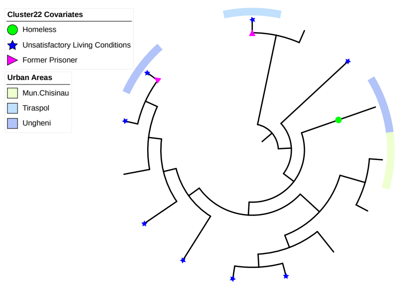


Cluster 22 with 14 isolates

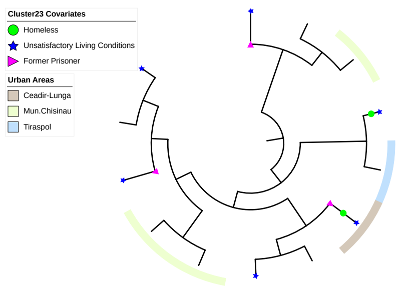


Cluster 23 with 13 isolates

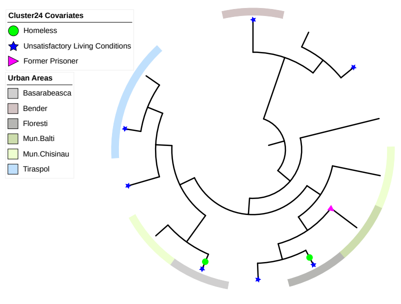


Cluster 24 with 13 isolates

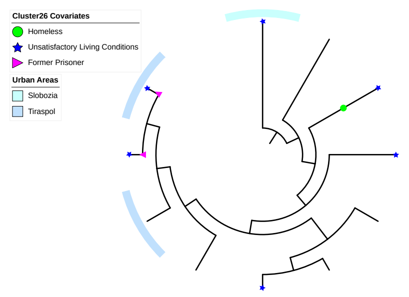


Cluster 26 with 11 isolates

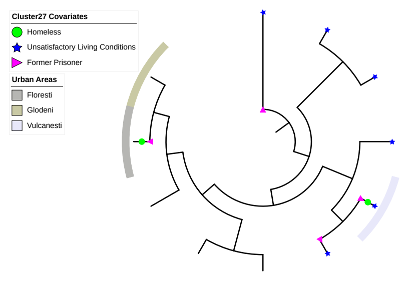


Cluster 27 with 11 isolates

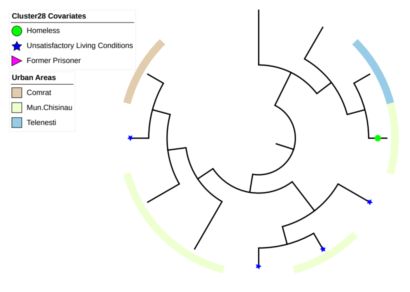


Cluster 28 with 11 isolates

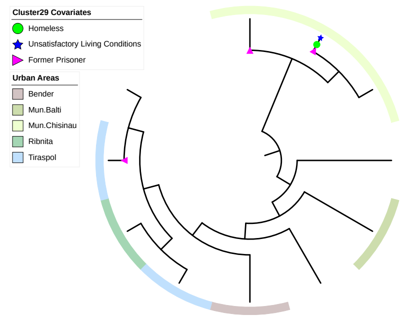


Cluster 29 with 11 isolates

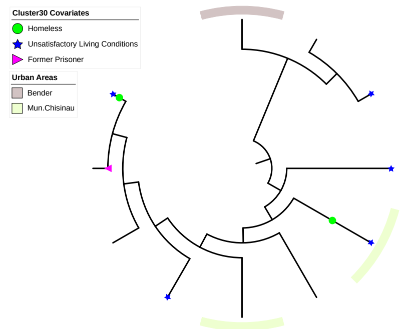


Cluster 30 with 11 isolates

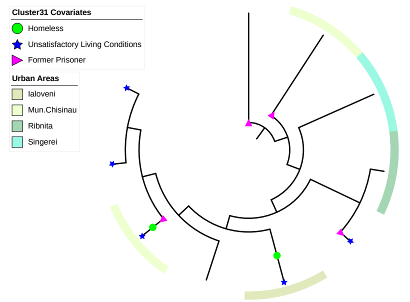


Cluster 31 with 10 isolates

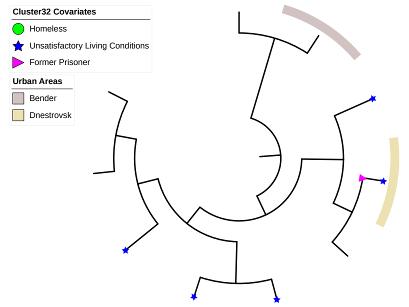


Cluster 32 with 10 isolates

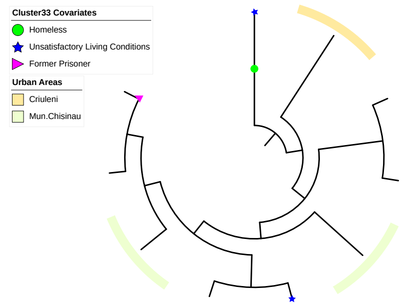


Cluster 33 with 10 isolates

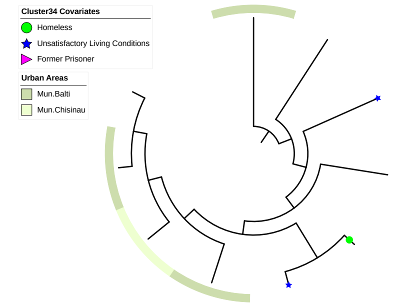


Cluster 34 with 10 isolates

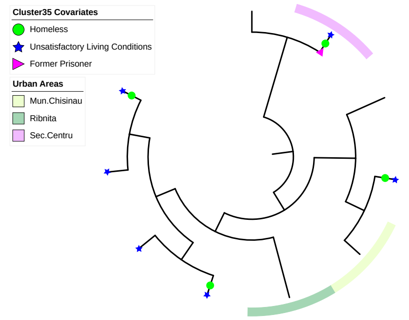


Cluster 35 with 10 isolates

We used a coalescent constant population model with a log normal [0,200] prior distribution. The optimal substitution model for individual clusters was selected on the basis of the Bayesian information criterion (BIC) score using Model-Finder. We adopted the BEAST2 correction for ascertainment bias, defining the number of non-variant A, C, G, and T sites for each cluster, which was manually added to the pre-processed file. We verified chain convergence, as well as good mixing, by calculating effective sample sizes (ESS) (greater than 200) for all parameters across each cluster using Tracer v1.7.1.

**Table S4:** Complete Coalescent Bayesian Skyline results of the sensitivity analysis using three different clock model settings (strict, log normal relaxed and exponential relaxed), and three clock rate estimates of the three large clades with specific resistant mutations.

| | | | | | | **Unfixed clock rate updated through the MCMC iterations** | | **Fixed lineage-specific clock rate based on previous estimates from literature** | | **Fixed clock rate based on prior BEAST analysis of the clades using a constant population** | |
|---|---|---|---|---|---|---|---|---|---|---|---|
| **Clades based on DR mutations** | **# of Taxa** | **Substitution Model** | **Method** | **Clock Rate Model** | **Clock Rate Distribution** | **tMRCA** | **Clock Rate (SNPs per site per year)** | **tMRCA** | **Clock Rate (SNPs per site per year)** | **tMRCA** | **Clock Rate (SNPs per site per year)** |
| Ural Clade 1 | 243 | TIM with unequal base frequencies | Coalescent Bayesian Skyline | Strict Clock | Log Normal | mean: 1968, 95% HPD interval: 1943 - 1988 | mean: 2.123 x $10^{-7}$, 95% HPD interval: 1.263 x $10^{-7}$ - 3.018 x $10^{-7}$ | mean: 1968, 95% HPD interval: 1944 - 1988 | mean: 2.132 x $10^{-7}$, 95% HPD interval: 1.267 x $10^{-7}$ - 2.998 x $10^{-7}$ | mean: 1967, 95% HPD interval: 1942 - 1988 | mean: 2.087 x $10^{-7}$, 95% HPD interval: 1.239 x $10^{-7}$ - 2.967 x $10^{-7}$ |
| | | | | Relaxed Clock Exponential | Log Normal | mean: 2002, 95% HPD interval: 1987 - 2013 | mean: 4.958 x $10^{-7}$, 95% HPD interval: 2.787 x $10^{-7}$ - 7.131 x $10^{-7}$ | mean: 2003, 95% HPD interval: 1988 - 2013 | mean: 4.971 x $10^{-7}$, 95% HPD interval: 2.901 x $10^{-7}$ - 7.043 x $10^{-7}$ | mean: 2003, 95% HPD interval: 1989 - 2012 | mean: 4.949 x $10^{-7}$, 95% HPD interval: 2.986 x $10^{-7}$ - 7.085 x $10^{-7}$ |
| | | | | Relaxed Clock Log Normal | Log Normal | mean: 1984, 95% HPD interval: 1961 - 2004 | mean: 2.803 x $10^{-7}$, 95% HPD interval: 1.626 x $10^{-7}$ - 3.998 x $10^{-7}$ | mean: 1984, 95% HPD interval: 1961 - 2003 | mean: 2.805 x $10^{-7}$, 95% HPD interval: 1.636 x $10^{-7}$ - 3.954 x $10^{-7}$ | mean: 1984, 95% HPD interval: 1961 - 2003 | mean: 2.796 x $10^{-7}$, 95% HPD interval: 1.624 x $10^{-7}$ - 3.996 x $10^{-7}$ |
| Beijing Clade 2 | 102 | TVM with equal base frequencies | Coalescent Bayesian Skyline | Strict Clock | Log Normal | mean: 2008, 95% HPD interval: 2001 - 2013 | mean: 2.88 x $10^{-7}$, 95% HPD interval: 1.605 x $10^{-7}$ - 4.243 x $10^{-7}$ | mean: 2007, 95% HPD interval: 2001 - 2013 | mean: 2.847 x $10^{-7}$, 95% HPD interval: 1.581 x $10^{-7}$ - 4.178 x $10^{-7}$ | mean: 2008, 95% HPD interval: 2001 - 2013 | mean: 2.886 x $10^{-7}$, 95% HPD interval: 1.642 x $10^{-7}$ - 4.184 x $10^{-7}$ |

| | | | | | | | | | | | |
|---|---|---|---|---|---|---|---|---|---|---|---|
| | | | | Relaxed Clock Exponential | Log Normal | mean: 2013, 95% HPD interval: 2011 - 2015 | mean: 7.502 x $10^{-7}$, 95% HPD interval: 3.76 x $10^{-7}$ - 1.155 x $10^{-6}$ | mean: 2013, 95% HPD interval: 2011 - 2015 | mean: 7.37 x $10^{-7}$, 95% HPD interval: 3.757 x $10^{-7}$ - 1.134 x $10^{-6}$ | mean: 2013, 95% HPD interval: 2011 - 2015 | mean: 7.307 x $10^{-7}$, 95% HPD interval: 3.626 x $10^{-7}$ - 1.14 x $10^{-6}$ |
| | | | | Relaxed Clock Log Normal | Log Normal | mean: 2013, 95% HPD interval: 2010 - 2015 | mean: 6.311 x $10^{-7}$, 95% HPD interval: 2.449 x $10^{-7}$ - 1.081 x $10^{-6}$ | mean: 2013, 95% HPD interval: 2010 - 2015 | mean: 6.248 x $10^{-7}$, 95% HPD interval: 2.445 x $10^{-7}$ - 1.087 x $10^{-6}$ | mean: 2013, 95% HPD interval: 2010 - 2015 | mean: 6.415 x $10^{-7}$, 95% HPD interval: 2.652 x $10^{-7}$ - 1.084 x $10^{-6}$ |
| Beijing Clade 3 | 121 | TVM with equal base frequencies | Coalescent Bayesian Skyline | Strict Clock | Log Normal | mean: 2002, 95% HPD interval: 1994 - 2008 | mean: 3.402 x $10^{-7}$, 95% HPD interval: 2.031 x $10^{-7}$ - 4.827 x $10^{-7}$ | mean: 2002, 95% HPD interval: 1994 - 2008 | mean: 3.368 x $10^{-7}$, 95% HPD interval: 1.986 x $10^{-7}$ - 4.776 x $10^{-7}$ | mean: 2002, 95% HPD interval: 1994 - 2008 | mean: 3.403 x $10^{-7}$, 95% HPD interval: 2.033 x $10^{-7}$ - 4.842 x $10^{-7}$ |
| | | | | Relaxed Clock Exponential | Log Normal | mean: 2009, 95% HPD interval: 2002 - 2014 | mean: 8.96 x $10^{-7}$, 95% HPD interval: 4.796 x $10^{-7}$ - 1.333 x $10^{-6}$ | mean: 2009, 95% HPD interval: 2003 - 2014 | mean: 8.891 x $10^{-7}$, 95% HPD interval: 4.812 x $10^{-7}$ - 1.331 x $10^{-6}$ | mean: 2009, 95% HPD interval: 2002 - 2014 | mean: 8.974 x $10^{-7}$, 95% HPD interval: 4.929 x $10^{-7}$ - 1.327 x $10^{-6}$ |
| | | | | Relaxed Clock Log Normal | Log Normal | mean: 2006, 95% HPD interval: 1999 - 2012 | mean: 5.037 x $10^{-7}$, 95% HPD interval: 2.743 x $10^{-7}$ - 7.487 x $10^{-7}$ | mean: 2006, 95% HPD interval: 1999 - 2012 | mean: 5.005 x $10^{-7}$, 95% HPD interval: 2.658 x $10^{-7}$ - 7.594 x $10^{-7}$ | mean: 2006, 95% HPD interval: 1999 - 2012 | mean: 5.031 x $10^{-7}$, 95% HPD interval: 2.721 x $10^{-7}$ - 7.635 x $10^{-7}$ |

*Bayesian Skyline plot analysis*
We used a coalescent Bayesian Skyline model to infer the events of *M. tuberculosis* population expansion to estimate the effective population size change through time in three large clades that were identified in the study population that contained individuals with specific drug resistance mutations. We used an uncorrelated log normal relaxed molecular clock and ran the MCMC algorithm for 250 million iterations, retaining every 25,000-th step from posterior, with the resulting log files analyzed using Tracer v1.7.1 for MCMC convergence and ESS values of greater than 200 across all parameters.

We also conducted a sensitivity analysis using three different clock model settings (strict, log normal relaxed, and exponential relaxed), and three clock rate estimates (**Fig. S6**): 1) an unfixed clock rate updated through the MCMC iterations, 2) a fixed lineage-specific clock rate based on previous estimates from the literature[21], and 3) a fixed clock rate based on prior BEAST analysis of the clades using a constant population.

**Fig. S6:** Coalescent Bayesian Skyline plots of the sensitivity analysis using three different clock model settings (strict, log normal relaxed and exponential relaxed), and three clock rate estimates of the three large clades with specific resistant mutations.

| **Clades based on DR mutations** | **Clock Rate Model** | **Unfixed clock rate updated through the MCMC iterations** | **Fixed lineage-specific clock rate based on previous estimates from literature** | **Fixed clock rate based on prior BEAST analysis of the clades using a constant population** |
|---|---|---|---|---|
| Ural Clade 1 | Strict Clock | 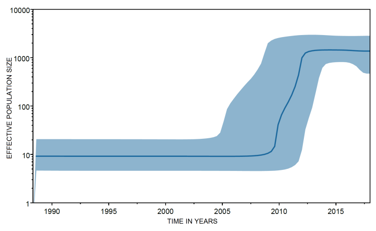 | 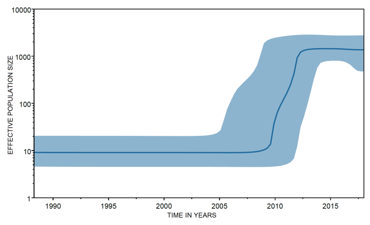 | 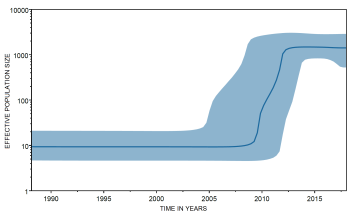 |
| | Exponential Relaxed Clock | 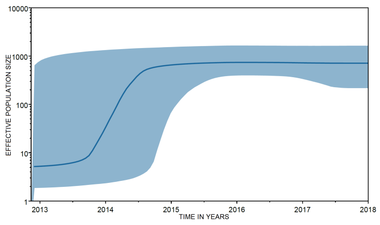 | 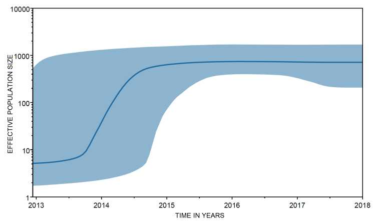 | 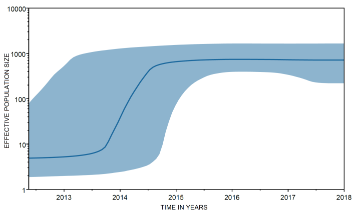 |
| | Log Normal Relaxed Clock | 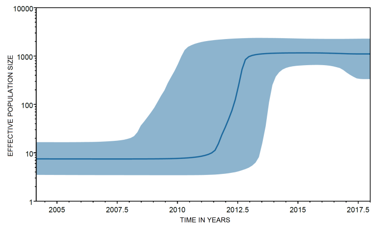 | 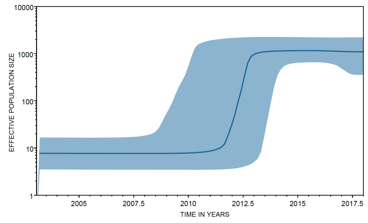 | 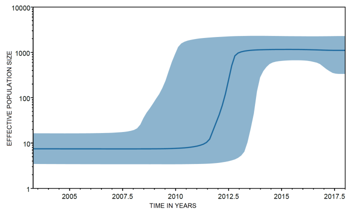 |

| | | | | |
|---|---|---|---|---|
| Beijing Clade 2 | Strict Clock | 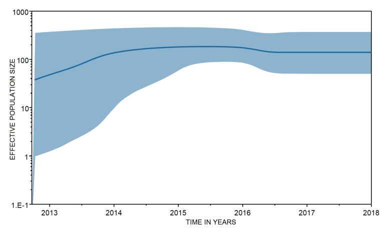 | 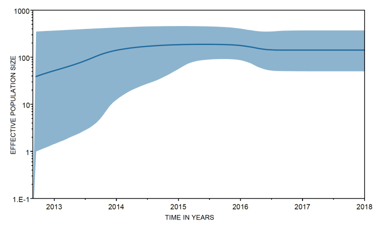 | 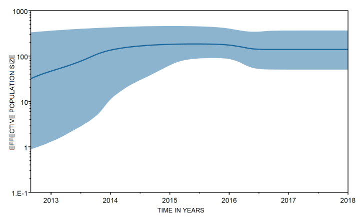 |
| | Exponential Relaxed Clock | 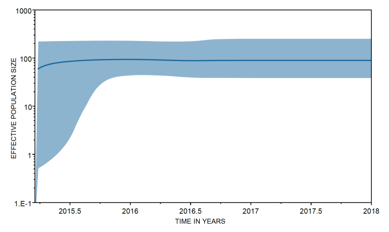 | 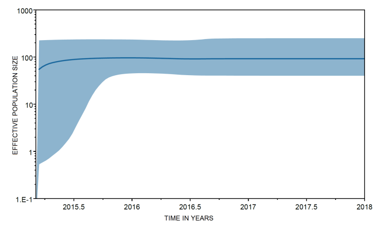 | 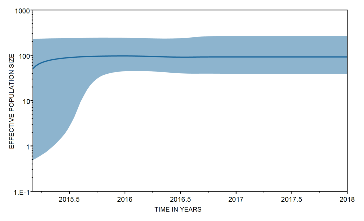 |
| | Log Normal Relaxed Clock | 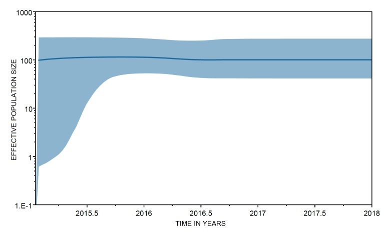 | 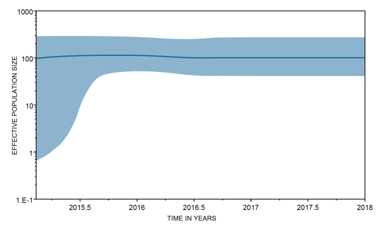 | 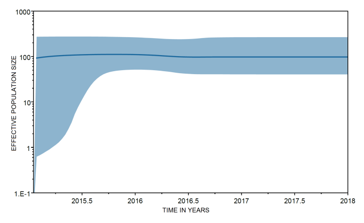 |
| Beijing Clade 3 | Strict Clock | 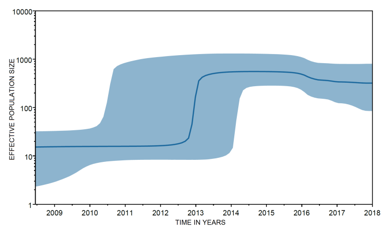 | 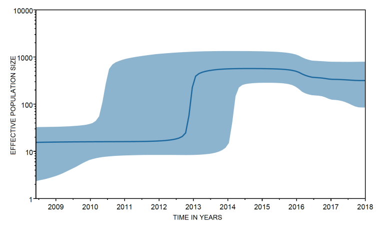 | 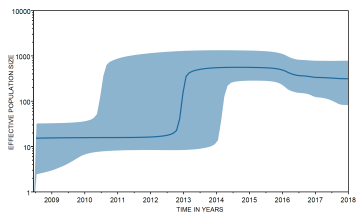 |
| | Exponential Relaxed Clock | 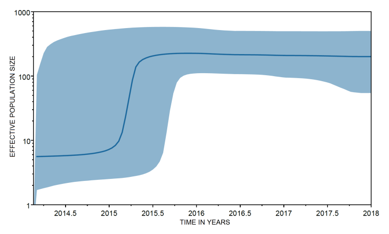 | 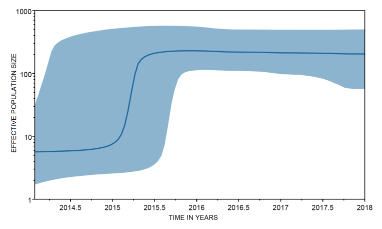 | 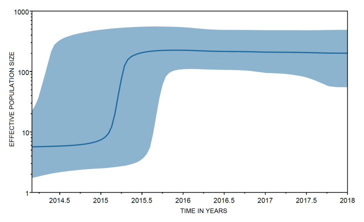 |
| | Log Normal Relaxed Clock | 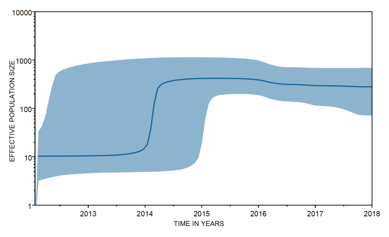 | 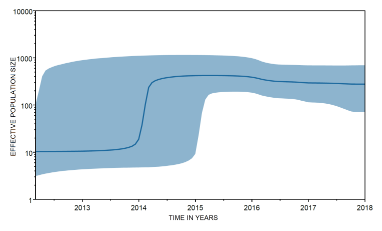 | 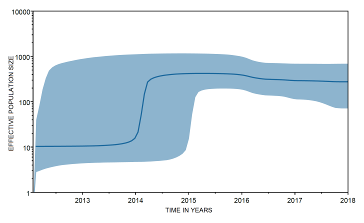 |

**S4. Spatial/genomic distance analysis**
We modeled the log-scaled patristic distance between each pair of cases as a function of geographic distance and other covariates such as

$$\ln(Y_{ij}) = \mathbf{x}_{ij}^{\mathrm{T}}\boldsymbol{\beta} + \left(\mathbf{z}_i + \mathbf{z}_j\right)^{\mathrm{T}}\boldsymbol{\gamma} + \epsilon_{ij}, i < j,$$

where $Y_{ij}$ is the patristic distance between cases i and j and $\epsilon_{ij} \sim \mathrm{N}(0, \sigma_\epsilon^2)$ are the independent, Gaussian distributed errors. We defined the expected value as a function of pair- and individual-level information, where $\mathbf{x}_{ij}$ includes covariates based on differences between the pair and $\mathbf{z}_i$ includes individual-level covariates.

We fit the regression model separately for each of the transmission clusters with at least 10 cases. For each analysis, we included a predictor if <10% of the values across the pairs were missing and if there were >4 pairs in each of the categorical variable levels, to ensure stable model fitting results. To better understand shared trends and variability in the estimated associations across genetic clusters, we then used the estimates and standard errors obtained from the first stage analyses within a Bayesian meta-analysis framework. The model for a single association is given as

$$\hat{\beta}_{jk} | \beta_{jk} \sim \mathrm{N}\left(\beta_{jk}, \hat{\sigma}_{jk}^2\right), k = 1, \ldots, 35,$$

$$\beta_{jk} \sim \mathrm{N}\left(\mu_{\beta_j}, \sigma_{\beta_j}^2\right)$$

where $\hat{\beta}_{jk}$ is the estimate obtained from the regression model fit to cluster k for covariate j, $\beta_{jk}$ represents the corresponding true but unobserved value, and $\hat{\sigma}_{jk}^2$ is the squared standard error of the estimate. We assumed that the true cluster-specific effects arise from a common Gaussian distribution with mean $\mu_{\beta_j}$ and variance $\sigma_{\beta_j}^2$, and estimate these parameters by giving them weakly informative prior distributions such that $\mu_{\beta_j} \sim \mathrm{N}(0, 100^2)$ and $\sigma_{\beta_j} \sim \mathrm{Uniform}(0, 100)$. By making inference on $\mu_{\beta_j}$ we determined if covariate j had a consistent impact when data were pooled across all clusters and uncertainty in the parameter estimates was correctly quantified. When reporting results from the second stage analysis, we present posterior means and 95% quantile-based credible intervals for $\exp\{\mu_{\beta_j}\}$ (i.e., the pooled effect on the relative risk (RR) scale). As a sensitivity analysis, we repeated these analyses modeling SNP distance (instead of patristic) using a similar Poisson regression framework.

For the spatial and genomic distance model, we performed a sensitivity analysis by modeling the SNP distance (instead of the patristic distance) between a pair of cases as a function of geographic distance and other covariates using a Poisson regression framework.

In **Table S4** we show the pooled RR inference for each of the effects. With respect to geographic distance, we see that two cases in the same locality have a 22% reduction in expected SNP distance compared to cases in different localities (RR: 0.78 (0.66, 0.99)). For these cases in different locality, as the distance between the localities increases by about 50 kilometers, the SNP distance between the pair increases by about 6% (RR: 1.06 (1.02, 1.09)). The only other significant effect is sex, where pairs comprised of two females tend to have larger SNP distances than male only (RR: 0.89, (0.81, 0.97)) or mixed (RR: 0.94 (0.90, 0.98)) pairs.

**Table S5:** Pooled Bayesian meta-analysis inference for each effect on the relative risk scale. Posterior means and 95% quantile-based credible intervals are presented.

| Effect | Estimate | 95% Credible Interval |
|---|---|---|
| Distance Between Centroids (50 km) | 1·06 | (1·02, 1·09) |
| Same Centroid (Yes vs. No) | 0·78 | (0·66, 0·99) |
| Date of Diagnosis Distance (1/2 year) | 1·01 | (1·00, 1·03) |
| Age Difference (10 years) | 1·01 | (0·99, 1·02) |
| Age (10 years) | 0·99 | (0·97, 1·00) |
| Household Contacts (1 person) | 1·00 | (0·98, 1·02) |
| Sex: | | |
| Mixed Pair vs. Both Female | 0·94 | (0·90, 0·98) |
| Both Male vs. Both Female | 0·89 | (0·81, 0·97) |
| Residence Location: | | |
| Mixed Pair vs. Both Not Urban | 1·06 | (0·99, 1·12) |
| Both Urban vs. Both Not Urban | 1·12 | (0·99, 1·26) |
| Housing: | | |
| Mixed Pair vs. Both Not Homeless | 1·03 | (0·87, 1·22) |
| Both Homeless vs. Both Not Homeless | 1·09 | (0·75, 1·57) |
| Working Status: | | |
| Mixed Pair vs. Both Unemployed | 1·05 | (0·97, 1·14) |
| Both Employed vs. Both Unemployed | 1·13 | (0·96, 1·33) |
| Education: | | |
| Mixed Pair vs. Both Secondary | 0·98 | (0·96, 1·01) |
| Both < Secondary vs. Both Secondary | 0·98 | (0·92, 1·05) |

### S5 Inference of direct transmission events

We identified direct transmission events between sampled hosts in large transmission clusters (⩾ ten cases, TreeCluster distance threshold 0.001 substitutions/site) by reconstructing transmission networks using TransPhylo.[24] This R package uses a Bayesian approach to reconstruct transmission networks from timed phylogenies, including sampled and un-sampled hosts, and allows for within-host diversity. We used a "multi-tree" method that simultaneously infers transmission trees from a selection of input phylogenetic trees while estimating a single value for shared model parameters. This accounts for uncertainty in the phylogenetic tree reconstruction.[24]

For each large transmission cluster (⩾ ten cases), we ran TransPhylo on 50 random trees drawn from a posterior selection of 10,000 timed trees produced in BEAST2, discarding the first 50% as burn-in, for a total of $10^5$ MCMC iterations. The parameter estimates for the offspring distribution, sampling density, and within-host coalescent rate were shared through multi-tree runs. For prior parameter values, the generation and sampling time prior parameters were represented by a gamma distribution with shape 1·3 and scale 3·33 (mean 4·3 years, SD 3·8 years), and 1·1 and 2·75 (mean 3·0 years, SD 2·9 years) respectively. While these prior distributions can have an impact on the resulting transmission inference, these values been used previously in *M. tuberculosis* transmission analyses[24] and allow the majority of transmission events occur within 2-3 years of the transmitting host being infected, but also accounts for the potential for long periods of asymptomatic latency. The offspring distribution was a negative binomial distribution with parameters $r$ = 1 (updated) and $p$ = 0.5 (fixed), and the within-host coalescent rate was fixed at 100/365. The sampling density (proportion of sampled cases) was updated through iterations and drawn from a beta distribution with strong informative priors α = 20 and ß = 8, reflecting the high proportion of culture-positive cases captured in this study. The resulting transmission trees were assessed to determine direct transmission events between sampled hosts with a posterior probability of ⩾0·5 (direct links between cases were found in more than half of the posterior transmission trees).

Reconstructing transmission networks in the 35 broad clusters using the multi-tree TransPhylo approach, we inferred 194 direct person-person transmission events. The relatively short study period allows for limited opportunities to capture transmission chains and pairs, and accordingly, a minority of clustered isolates were predicted to be involved in direct transmission events in at least half the posterior transmission trees (338/1000, 33.8%). Nonetheless, the identification of these direct transmission events supports evidence of recent, local transmission between sampled individuals in the region. We found no significant factors that were associated with inclusion in these direct transmission events compared to other clustered individuals, though there was some evidence for an increased likelihood of transmission linkage between hosts in the Transnistria region compared to the rest of Moldova (OR 1·42, $P$ = 0·02).
